# *Toxoplasma gondii* genotyping in 2,467 human toxoplasmosis cases from France and other geographical origins: genetic diversity and clinical associations

**DOI:** 10.64898/2026.10.26.26366474

**Authors:** Etienne Herault, Marion Vergonjeanne, Karine Passebosc-Faure, Thibaut Gelle, Marie-Fleur Durieux, Farid Boumediene, Jean-Benjamin Murat, Rose Anne Lavergne, Françoise Gay-Andrieu, Naima Dahane, Judith Fillaux, Feriel Touafek, Luc Paris, Marie-Pierre Brenier-Pinchart, Cécile Garnaud, Maude Lévêque, Yvon Sterkers, Christelle Pomares, Nicoles Ferret, Florence Robert-Gangneux, Hélène Guégan, Denis Filisetti, Odile Villard, Anne Sophie Deleplancque, Frédéric Dalle, Coralie L’Ollivier, Julie Bonhomme, Bénédicte Langlois, Nathalie Van Langendonck, Claire Hoffmann, Gilles Gargala, Isabelle Accoceberry, Juliette Guitard, Martine Wallon, Estelle Perraud, Sandrine Houzé, Anne Totet, Céline Damiani, Céline Nourrisson, Emeline Scherer, Frédéric Grenouillet, S. Hamane, Pierre Flori, Anne Debourgogne, Nicole Desbois-Nogard, Thomas Nicol, Marwan Haboub, Lorra Monpierre, Sophie Brun, Laura Verdurme, Marie-Elisabeth Bougnoux, Nicolas Traversier, Françoise Botterel-Chartier, Elise Benard, Anaïs Soares, Christelle Vauloup-Fellous, Isabelle Villena, Daniel Ajzenberg, Aurélien Mercier, Marie-Laure Dardé, Hélène Yera, Lokman Galal

## Abstract

**Background:** *Toxoplasma gondii* exhibits marked variation in genetic diversity, yet the relationships between parasite genotype and clinical presentation in humans remain poorly understood. We investigated the diversity of *T. gondii* strains infecting humans and their associations with different forms of toxoplasmosis.

**Methodology/Principal Findings:** We retrospectively analyzed 2,467 *T. gondii-*positive human clinical cases collected in France through a nationwide network of hospital parasitology diagnostic units over more than four decades. Cases included patients receiving care in mainland France, including residents, travelers, and immigrants, as well as patients managed in French overseas territories in South America, the Indian Ocean, and Oceania. Parasite genotyping was performed using a standardized panel of 15 microsatellite markers. Complete genotypes were obtained for 1,269 cases, revealing 13 distinct genetic groups, including clonal lineages and non-clonal populations, alongside genotypes that could not be assigned to any recognized group. Type II strains predominated overall (82.6%) and were particularly frequent in congenital toxoplasmosis (869/955 cases, 91.0%). Despite being predominantly associated with infraclinical or mild congenital infections, type II strains were also responsible for severe congenital infections following maternal infection during all three trimesters of pregnancy, as well as acquired ocular toxoplasmosis. Haplogroup 16, encompassing strains previously assigned to multiple PCR-RFLP genotypes, showed an association with severe congenital toxoplasmosis, with six of seven cases presenting severe manifestations. Type I strains were remarkably rare (n = 5) and showed no signs of increased clinical severity compared to type II. Amazonian strains, another genetically diverse group, were identified almost exclusively in acquired toxoplasmosis in immunocompetent patients, all of them severe, in keeping with previous reports of the exceptional pathogenicity of this genetic group. In immunodeficient patients, Africa 1 strains (equivalent to ToxoDB#6) were more frequently associated with isolated cerebral toxoplasmosis (20/25 cases, 80%) than type II and type III strains. Overall, the results support that while a given *T. gondii* genetic group may be associated with a broad spectrum of clinical manifestations, significant differences in disease presentation exist between genetic groups.

**Conclusions/Significance:** This study constitutes the largest collection of human *T. gondii* genotypes linked to clinical data assembled to date and provides new insights, at a global level, into the relationships between parasite genetic diversity and clinical manifestations of toxoplasmosis. Our findings suggest a role for parasite genotype in shaping disease presentation, even in the context of strong host-related determinants such as immunodeficiency or gestational timing of infection. These results also highlight the value of long-term surveillance networks for understanding the epidemiology and pathogenicity of *T. gondii* on a global scale.

## 1. Introduction

Toxoplasmosis is a ubiquitous parasitic infection caused by *Toxoplasma*, a zoonotic protozoan that infects warm-blooded animals, including humans [1]. It is estimated to affect approximately one third of the world’s population [2]. To date, only one species, *Toxoplasma gondii*, is recognized as responsible for human toxoplasmosis; however, the disease presents with highly diverse clinical manifestations. Several factors influence clinical presentation, including host immune status [3], parasite inoculum [4,5], parasite genetic diversity [6–8], age, ethnicity of the host [9,10], and gestational trimester in congenital toxoplasmosis [11].

Knowledge of *T. gondii* diversity has considerably expanded over the past four decades. Initial studies [4,12,13] identified three clonal lineages (types I, II, and III). Subsequently, broader sampling encompassing a greater diversity of host species and geographic origins, together with the development of multilocus genotyping approaches, revealed a much broader global genetic diversity. These approaches primarily relied on microsatellite (MS) [14,15] and restriction fragment length polymorphism (RFLP) markers [4,16].

These genetically diverse strains are associated with markedly different phenotypes in both mice and humans [17,18]. The frequent use of mouse bioassays for strain isolation, together with the ease of performing experimental infections in mice, has enabled detailed characterization of the relationship between *T. gondii* genotypes and toxoplasmosis severity [17]. In mice, strains are generally classified as non-virulent when infection remains asymptomatic, virulent when it results in acute lethal infection, and intermediate when both phenotypes are observed. In humans, however, the situation is far more complex. In addition to the existence of several clinical forms of toxoplasmosis, the spectrum and severity of clinical manifestations within each form are highly heterogeneous, reflecting the influence of multiple factors beyond the intrinsic characteristics of the parasite [10,18]. Furthermore, reports describing *T. gondii* genotypes in clinical samples remain sparse, are often limited to individual case reports or small case series, and rely on heterogeneous genotyping methods and genotypes designation schemes [19], including the still frequent use of genotyping techniques with insufficient discriminatory power to distinguish between *T. gondii* lineages and populations (e.g., single-locus marker genotyping) [20–25]. Consequently, associations between parasite genotypes and clinical manifestations are still poorly understood and remain difficult to synthesize and harmonize.

In France, a nationwide network of hospital parasitology diagnostic units has developed for more than three decades and has been constituted in a National Reference Centre for Toxoplasmosis (NRCT) since 2006, that has among several missions the molecular surveillance of *T. gondii* strains in pregnant women undergoing follow-up and in patients diagnosed with toxoplasmosis throughout France through genotyping approaches (https://cnrtoxoplasmose.chu-reims.fr/?lang=en). Until 2023, the NRCT and the associated network of parasitology diagnostic laboratories collected 2,467 toxoplasmosis cases reported in France, including cases from mainland France, French overseas territories, travellers, and immigrants. The corresponding strains originated predominantly from mainland France, but also from other European countries, Africa, Asia, Oceania, and South America.

In the present study, we performed a retrospective analysis of the genotypic and genetic diversity of *T. gondii* strains collected for the surveillance of toxoplasmosis in France, with the aim of evaluating associations between *T. gondii* genotypes and the clinical presentations of patients with toxoplasmosis.

## 2. Material and methods

### 2.1. Sample and Data collection

In France, epidemiological surveillance of human toxoplasmosis has officially been coordinated by the NRCT since 2006, following the implementation of the national online toxoplasmosis reporting system, ToxoSurv [26]. Surveillance primarily focuses on cases of congenital toxoplasmosis (CT) identified through routine prenatal screening, for which systematic monitoring was established to reduce the risk of undiagnosed maternal infections. However, surveillance also includes other clinical forms of toxoplasmosis, such as acquired toxoplasmosis (as opposed to congenital) in immunocompetent (ATIC) or immunodeficient patients (ATID) and isolated ocular toxoplasmosis (OT). Within this framework, the Laboratory of Parasitology of the Limoges University Hospital is responsible for molecular surveillance through the genotyping of *T. gondii* directly from DNA extracts from clinical samples or from strains isolated from patients. Nevertheless, this genotyping activity had already begun several years before the establishment of the NRCT, through a nationwide network of hospital correspondents sending samples to the Limoges laboratory for genotyping. Samples were sent by parasitology diagnostic laboratories in hospitals across mainland France, as well as from French overseas territories in Latin America and the Caribbean (French Guiana, Martinique, and Guadeloupe), in the Indian Ocean region (Réunion Island) and Oceania (French Polynesia and New Caledonia).

Samples consist either of DNA extracted from pathological products using the DNA extraction protocols routinely implemented in the participating hospital laboratories (Ct < 33 with quantitative PCR), or of *T. gondii* strains isolated by mouse bioassay from these pathological products. In the latter case, isolates are sent by NRCT network correspondents in the form of infected mouse tissues (brain or ascitic fluid). DNA extraction from these materials is subsequently performed at the Limoges laboratory using the QIAamp DNA Minikit® (QIAGEN®, Germany).

Correspondents also provide epidemiological (country of residency or country of origin, travel history outside France, potential sources of contamination) and clinical data (immune status and main clinical manifestations). In the present study, samples sent between January 2006 and December 2023 by the correspondents of the NRCT had their associated data entered through ToxoSurv. In addition, earlier cases referred to the Limoges laboratory before 2006 were also incorporated into the analysis, with the same types of epidemiological data available. Overall, the dataset comprised 1,975 toxoplasmosis cases collected from 2006 onwards, 433 cases collected before 2006, and 59 cases for which the year of sampling could not be retrieved.

### 2.2. Ethical statement

The analysis of biological samples obtained through standard clinical practices was classified as non-interventional research in accordance with Article L1221-1.1 of the French Public Health Code, requiring only the absence of patient objection at the time of sample collection (as per Article L1211-2 of the Public Health Code). At participating centers, mothers and patients were informed about the anonymized reporting of CT cases, the collection of clinical and epidemiological data, and the study of T. gondii isolates from different clinical forms of toxoplasmosis. Mothers and patients had the right to refuse participation in this survey. All collected data were anonymized before samples were sent to the Limoges laboratory. Anonymized clinical and epidemiological data associated with *T. gondii* strains collected through the French NRCT were reported by the 37 participating specialized laboratories via a dedicated online data-entry system (http://cnrtoxoplasmose.chu-reims.fr), using the specifically developed ToxoSurv software (EpiConcept). The software and associated data collection procedures were approved by the French National Data Protection Authority (Commission Nationale de l’Informatique et des Libertés, CNIL; approval reference DR-2016-186).

### 2.3. Clinical classification of toxoplasmosis cases

All reported toxoplasmosis cases were classified into four main categories according to their clinical presentation: CT, ATID, ATIC and OT. Within each group, additional subclassifications were defined according to specific clinical criteria (Table 1). For CT specifically, cases in which newborns were initially suspected of CT (e.g., positive placental screening at birth) but for whom follow-up ultimately excluded mother-to-child transmission were classified as no congenital toxoplasmosis (no-CT). The definition of CT cases was based on the classification by Lebech *et al*. [27].

**Table 1.** Definitions of clinical categories and clinical forms within each category.

| Main categories | Clinical form | Definition |
| --- | --- | --- |
| **Congenital toxoplasmosis (CT)** | Infraclinic CT | Absence of clinical signs at birth |
|  | Mild CT | Isolated cerebral calcifications and/or peripheral retinochoroidal lesions at birth |
|  | Severe ocular CT | Macular retinochoroidal lesions at birth |
|  | Severe CT | Still birth, *in utero* death, perinatal death, antenatal echographic signs resulting in medical abortion, ventricular dilatations or microcephaly at birth |
|  | CT with no or insufficient data | Absence of antenatal echographic signs, but missing information at birth<br>Missing information on clinical signs at birth |
| **Systemic or localized acquired toxoplasmosis in immunodeficient patients (ATID)** | Cerebral ATID | Isolated cerebral localisation, highlighted by clinical, iconographic and/or biological, defined by positive PCR in brain or spinal cord tissue or CerebroSpinal Fluid |
|  | Disseminated ATID | Extracerebral involvement (mainly lungs), often with positive PCR in blood or bone-marrow. Cerebral lesions may be associated. |
|  | Paucisymptomatic or asymptomatic ATID | Isolated fever or fever associated with lymphadenopathies or without any symptoms other than isolate ocular localisation |
| **Systemic or localized acquired toxoplasmosis in immunocompetent patients (ATIC)** | Paucisymptomatic ATIC | Lymphadenopathies, fever, possible association with hepatomegaly and/or splenomegaly, cutaneous rash |
|  | Severe ATIC | Disseminated forms, usually with pulmonary involvement, and/or other organ or tissue involvement other than isolate ocular localisation (myocarditis, pericarditis, polymyositis, ascites...) |
| **Isolated ocular toxoplasmosis (OT)** | Non-severe OT | Ocular toxoplasmosis cases showing no features suggestive of severe ocular disease, such as extensive inflammation (e.g., panuveitis), bilateral involvement, recurrent/reactivated disease, or incomplete clinical improvement. |
|  | Severe OT | Ocular toxoplasmosis cases presenting at least one feature suggestive of severe ocular disease, including extensive inflammation (e.g., panuveitis), bilateral involvement, recurrent/reactivated disease, or incomplete clinical improvement. |
|  | OT with insufficient clinical data | Ocular toxoplasmosis cases lacking clinical information to assess ocular disease severity. |

### 2.4. Geographical classification of toxoplasmosis cases

Geographical classification was defined based on the place of acquisition of infection. Classification followed the World Bank’s *Country and Lending Groups 2025* regional framework (East Asia and the Pacific; Europe and Central Asia; Latin America and the Caribbean; Middle East and North Africa; North America; South Asia; Sub-Saharan Africa), ensuring standardized geographic grouping. French Guiana, Guadeloupe, Martinique, Reunion Island and Mayotte are not listed separately in this framework but fall within France; they were assigned to the region of their geographical location rather than to Europe and Central Asia. French Polynesia and New Caledonia are listed separately and were classified accordingly.

### 2.5. *T. gondii* genotyping and genotype classification

Genotyping of samples within the framework of NRCT activities at the Limoges laboratory is based on multilocus analysis of MS markers. This MS typing method, originally developed at the Limoges laboratory, has evolved over time with regard to the number of markers used: from 5 MS markers between 2000 and 2006 [28], to 6 MS markers between 2006 and 2009 [29], and finally to a multiplex panel of 15 MS markers after 2009 [15,30]. To ensure methodological consistency and comparability over time, samples initially genotyped using the 5-or 6-marker panels were progressively reanalysed using the 15-marker panel.

Among the 15 MS markers, 8 are considered typing markers, and the combination of alleles at these loci allows classification of strains into clonal lineages (e.g., clonal types I, II, and III, Africa 1–4, and Caribbean 1–3). For type II and Africa 4, a difference of one repeat unit (2 bp) at a single typing marker was considered compatible with assignment to the respective lineage. This criterion accounts for the greater microsatellite variability of type II and Africa 4, two lineages whose longer evolutionary history, suggested by whole-genome analyses [31], may have allowed greater accumulation of mutations compared with other clonal lineages.

However, some genotypes belong to partially clonal or non-clonal *T. gondii* populations, which display allele polymorphism at typing loci and therefore lack a unique allelic signature. Some of these genotypes could not be assigned to recognized clonal lineages based on their typing profiles but corresponded to genetic groups previously characterized by whole-genome sequencing, namely haplogroup 16 and the Amazonian group [31,32]. Strains previously assigned to these groups by whole-genome analyses retained their classification. A neighbour-joining tree was built from all 15 MS markers using Bruvo’s genetic distance and the BRUVO.BOOT function of the poppr package in R. Additional genotypes were assigned to haplogroup 16 or to the Amazonian group when they clustered with the corresponding WGS-characterized strains in a branch separated from all other genotypes in the neighbour-joining tree constructed from the 15 MS markers. Genotypes that could not be assigned according to these criteria were classified as unclassified. Because this category is heterogeneous by construction and does not correspond to a genetic group, it was described but not included in comparisons between genetic groups.

### 2.6. Associations between *T. gondii* genetic groups and clinical presentations

The evaluation of associations between *T. gondii* genetic groups and the clinical presentations of patients with toxoplasmosis included only strains successfully genotyped at all 15 MS markers. Unless otherwise stated, proportions of clinical forms were calculated among cases with documented clinical data. First, the proportions of type II genotypes versus non-type II genotypes were compared pairwise between the four clinical categories (CT, ATIC, ATID, and OT) using Chi-squared or Fisher’s exact tests, as appropriate. Odds ratios (ORs) and corresponding 95% confidence intervals (95% CIs) were obtained by conditional maximum likelihood. Both uncorrected p-values and p-values after Bonferroni correction for the number of comparisons within each family of tests are reported. Then, within each clinical category, multiple correspondence analysis (MCA) were performed as a hypothesis-generating approach to explore relationships between *T. gondii* genetic groups and clinical forms, using the FactoMineR package in R (version 4.3.0; Vienna, Austria). MCA is an extension of correspondence analysis (CA) and a generalization of principal component analysis (PCA). This multivariate analysis technique allows the investigation of potential relationships between multiple categorical variables [33–36]. MCA results are presented graphically in two-dimensional graphs, defined by the dimensions permitting the largest proportion of inertia. Because complete disjunctive coding mechanically deflates the raw proportions of inertia of an MCA, which are therefore not comparable to the proportions of variance of a PCA, adjusted inertias are reported. Only genetic groups represented by at least three toxoplasmosis cases were included. The analysis also accounted for the geographic origin of infection and for category-specific variables, such as the gestational trimester in CT cases and the type of immunosuppression in ATID cases. In some CT cases, it was not possible to determine the precise date of maternal infection because of late performance of the initial serological test or irregular monthly serological monitoring; so infections occurring between 0 and 6 weeks of gestation were classified as periconceptional, while those falling between two trimesters were classified as “1st–2nd trimester” and “2nd–3rd trimester”. In a few CT cases, maternal infection was acute but preconceptional or the mother had a chronic infection prior to pregnancy. All such CT cases where the date of maternal infection could not be determined during pregnancy or who were infected prior to pregnancy were excluded from the MCA and pairewise comparisons.. Hypotheses generated by the MCA were subsequently evaluated using pairwise Chi-squared or Fisher’s exact tests, as appropriate. Because the same dataset generated and tested these hypotheses, the corresponding p-values are reported as exploratory.

## 3. Results

### 3.1. Clinical cases and clinical samples

The dataset comprised 2,709 samples from 2,467 human toxoplasmosis cases recorded between 1962 and 2023 (S1 table), including 433 cases prior to 2006 (17.6%), 1,975 cases from 2006 onwards (80.1%) and 59 cases for which the year of sampling could not be retrieved (2.4%). Among the 2,467 cases, 1,062 (43.0%) were documented by a strain isolated by mouse bioassay and 1,405 (57.0%) by a primary clinical specimen only. Among the 2,467 cases, information on the main clinical category was available for 2,444 cases (99.1%), while detailed clinical presentation data were available for 2,149 cases (87.1%). Information on the place of acquisition of infection was available for 1,978 cases (80.2%). Overall, 2,244 cases (91.0%) corresponded to samples obtained from a single clinical matrix, whereas the remaining cases involved more than one matrix (e.g., brain biopsy and cerebrospinal fluid [CSF], or placenta and amniotic fluid [AF]). The matrices were highly diverse and included, in decreasing order of frequency, placentas (n = 817), amniotic fluids samples (AF; n = 623), aqueous humors (AH; n = 300), blood or serum samples (n = 293), CSF samples (n = 208), brain biopsies (n = 136), bronchoalveolar lavage samples (BAL; n = 100), umbilical cord blood samples (n = 71), vitreous humors (VH; n = 38), various fetal tissues (n = 41), lymph nodes (n = 22), bone marrow samples (n = 17), and 43 samples originating from 18 other matrix types.

### 3.2. *T. gondii* genotyping

Among the 2,467 cases, complete 15 MS genotyping was successfully obtained for 1,269 cases (51.4%). Successful 15 MS genotyping was achieved far more frequently from isolated strains (927/1,062; 87.3%) than from primary clinical specimens (342/1,405; 24.3%). Genotyping success also varied markedly across clinical categories, from 69.4% in congenital toxoplasmosis (CT) to 7.1% in isolated ocular toxoplasmosis (OT), and declined over time, from 84.8% before 2000 to 37.7% in 2016-2023, in parallel with the shift from strain isolation to direct genotyping from clinical specimens (S2 Table). Within clinical categories, genotyping success was also higher for the more severe clinical forms in CT and in acquired toxoplasmosis of both immunodeficient (ATID) and immunocompetent (ATIC) patients, but not in ocular toxoplasmosis (OT) (S2 table). Cases with complete genotyping data 955 cases of CT(75.3%), 9 cases for no-CT(0.7%), 238 cases of ATID (18.8%), 42 cases of ATIC (3.3%), and 23 cases of isolated OT(1.8%). Regarding the place of acquisition of infection, 1,102 cases were in Europe and Central Asia (86.8%), 70 in Latin America and the Caribbean (5.5%), 55 in sub-Saharan Africa (4.3%), 9 in East Asia and the Pacific (0.7%), 1 in the Middle East and North Africa (0.1%), and 32 were classified as undefined (2.5%) (S3 Table).

### 3.3. *T. gondii* genotypes classification

The neighbor-joining tree generated from the 1,269 *T. gondii* strains successfully genotyped at all 15 MS markers (Fig. 1) showed a clear separation between genetic groups. Genotypes belonging to the same genetic group generally clustered together, with only a few exceptions. Strains previously assigned to haplogroup 16 by whole-genome analyses formed a distinct cluster, together with additional genotypes that were consequently assigned to this group. In contrast, WGS-characterized Amazonian strains did not form a distinct cluster and could not be reliably distinguished from other genotypes on the basis of their MS profiles. The Amazonian designation was therefore restricted to strains previously assigned to this group by whole-genome analyses and to three strains sharing a strictly identical 15-marker profile with one of them (TgH18010A and TgH18014A, identical to TgH18008A; TgH18070A, identical to TgH18066A) (S1 table). The pattern highlights the exceptionally high genetic diversity of this population and indicates that 15 microsatellite markers do not resolve its boundaries. Overall, based on the combination of alleles at the eight typing markers and divergence tree-based clustering analyses,1,048 strains were classified as type II (82.6%), 38 as type III (3.0%), 34 as Africa 1 (2.7%), 16 as Amazonian (1.3%), 10 as haplogroup 16 (0.8%), 9 as Caribbean 1 (0.7%), 6 each as Caribbean 3 and Africa 4 (0.5% each), 5 each as type I, Africa 2 and Polynesian 1 (0.4% each), and 4 each as Caribbean 2 and Mascarene 1 (0.3% each). The remaining 79 strains (6.2%) could not be assigned to any recognized genetic group and were classified as unclassified.

**Fig. 1.**
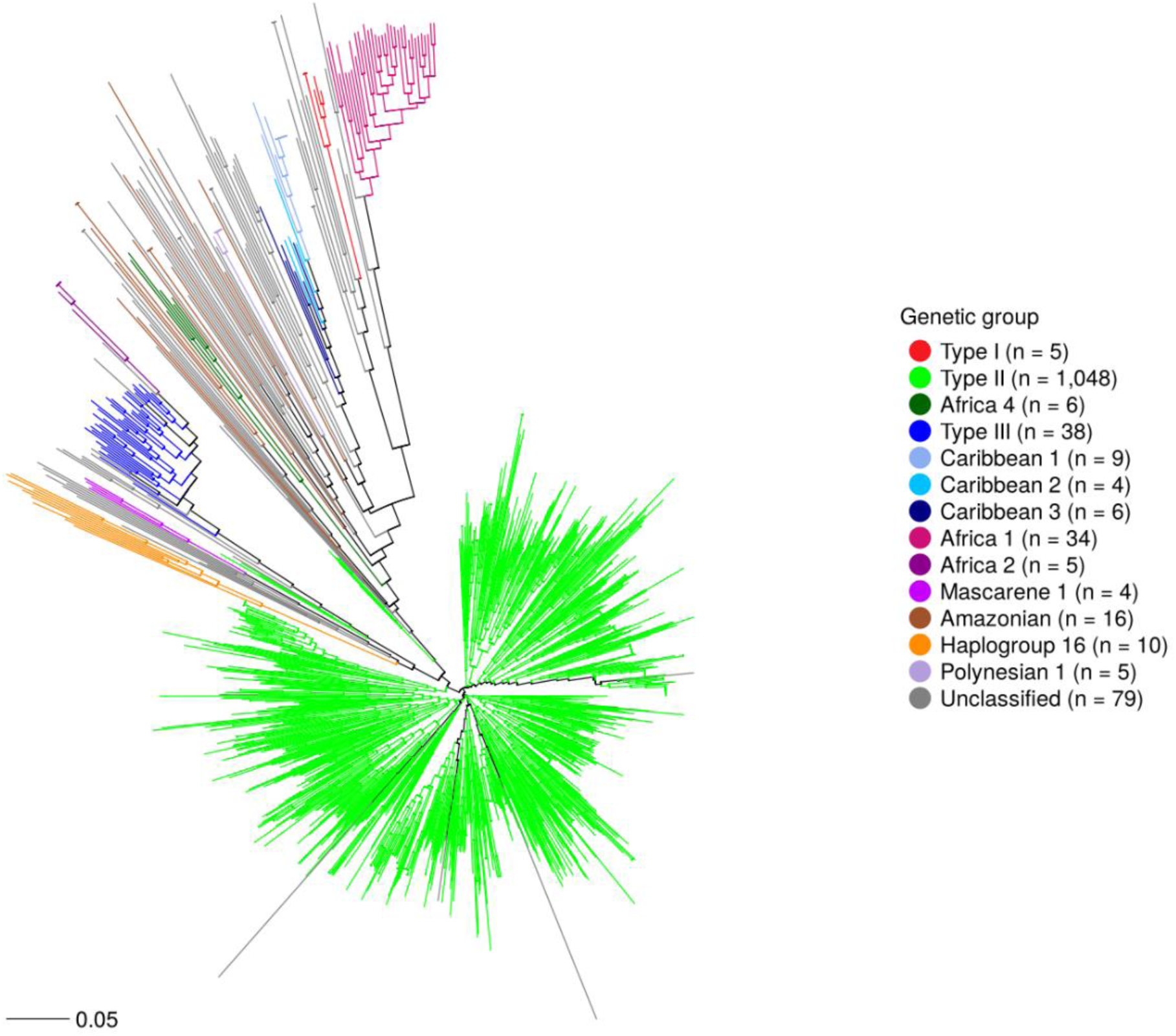
Neighbor-joining divergence tree of 1,269 *Toxoplasma gondii* strains constructed from calculated Bruvo distances using 15 microsatellite loci.

**Fig. 2.**
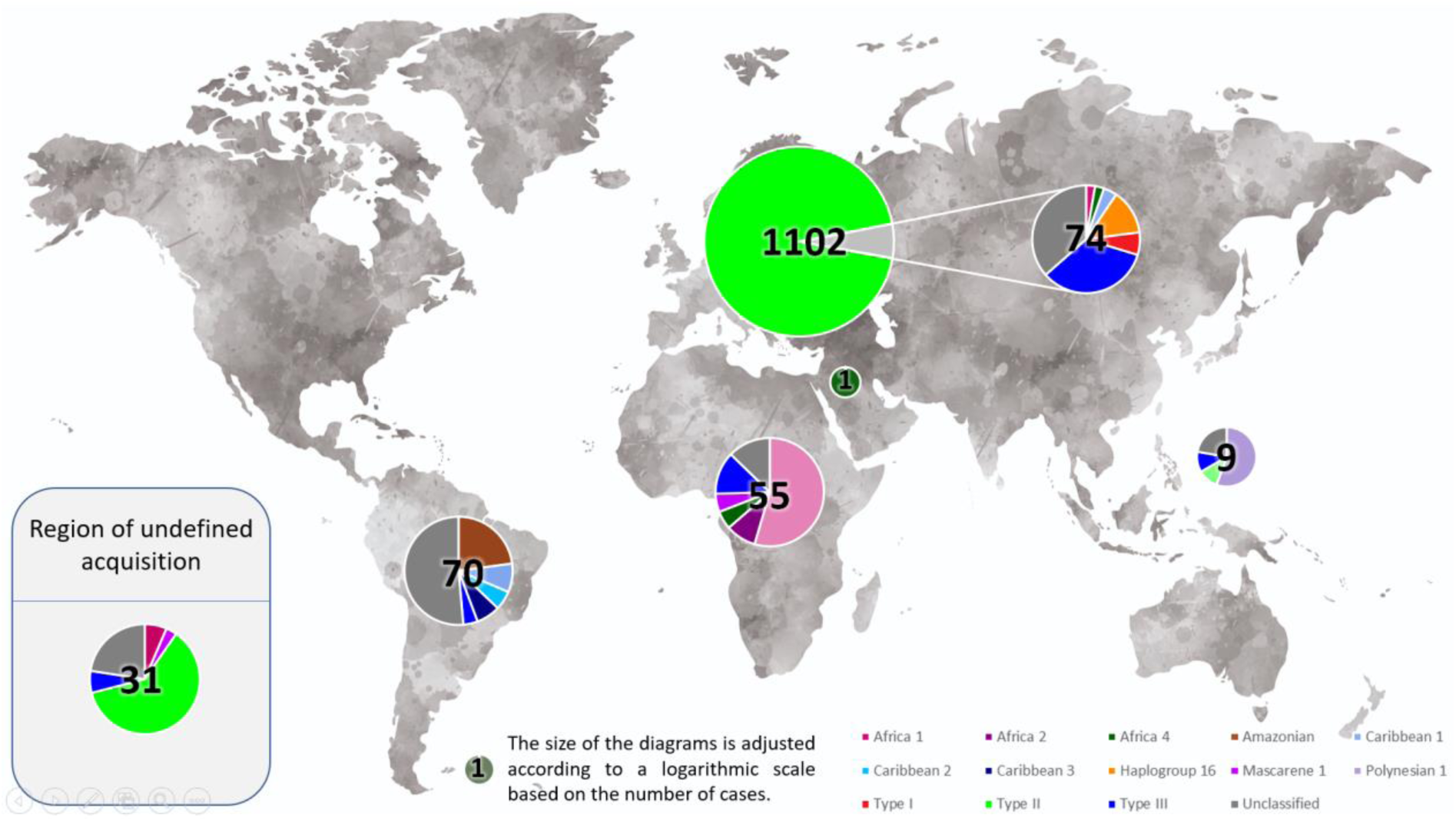
Geographic distribution of *Toxoplasma gondii* genotypes among human toxoplasmosis cases with 15 microsatellite loci.

### 3.4. Geographical classification of toxoplasmosis cases

Type II was overwhelmingly associated with infections acquired within the Europe and Central Asia region (1,028/1,048 strains, 98.1%) (S3 Table). Conversely, type II accounted for 93.3% (1,028/1,102) of all infections acquired in this region. Europe and Central Asia was also the place of acquisition for all haplogroup 16 strains (10/10, 100%), for most type III strains (25/38, 65.8%), for all five type I strains (100%) and for 27 of the 79 unclassified genotypes (34.2%). Of note, reported consumption of imported horse meat was documented in 13 of the complete genotyped cases, involving 10 unclassified genotypes, 2 type II strains and 1 haplogroup 16 strain, all acquired in Europe. While haplogroup 16 was exclusively associated with this region, type III strains showed a broader geographic distribution, with cases also attributed to sub-Saharan Africa (7/38, 18.4%), Latin America and the Caribbean (3/38, 7.9%), and East Asia and Pacific (1/38, 2.6%).

Other genetic groups showed strong associations with geographic regions outside Europe and Central Asia. Most Africa 1 (30/34, 88.2%) and all Africa 2 strains (5/5, 100%) were associated with sub-Saharan Africa, as were three of the four Mascarene 1 strains (75.0%), all identified in Reunion Island, a French overseas territory of the Indian Ocean. Caribbean lineages 1, 2, and 3 were primarily associated with the French overseas territories of Martinique, Guadeloupe, and French Guiana in the Latin America and Caribbean region, with frequencies of 66.7% for Caribbean 1of 100% for Caribbean 2 and of 83.3% for Caribbean 3. Similarly, all 16 Amazonian strains were acquired in Latin America and the Caribbean (100%), 13 of them in French Guiana and 3 in Suriname, while the Polynesian 1 lineage was exclusively associated with French Polynesia in the East Asia and Pacific region (100%). Unclassified genotypes were the most geographically heterogeneous group, with 36 of 79 strains (45.6%) acquired in Latin America and the Caribbean.

### 3.5. Associations between *T. gondii* genetic groups and clinical presentations

Although type II strains predominated overall, their distribution varied considerably across clinical categories. They accounted for 91.0% (869/955) of CT cases, 65.5% (156/238) of ATID cases and 60.9% (14/23) of OT cases, but only 7.1% (3/42) of ATIC cases (S3 Table). All pairwise comparisons were statistically significant after Bonferroni correction except that between ATID and OT: CT versus ATID (OR = 5.30; 95% CI 3.69-7.62; corrected p < 0.0001), CT versus ATIC (OR = 130.0; 95% CI 40.1-671.2; corrected p < 0.0001), CT versus OT (OR = 6.47; 95% CI 2.40-16.6; corrected p = 0.0008), ATID versus ATIC (OR = 24.5; 95% CI 7.43-127.5; corrected p < 0.0001), ATIC versus OT (OR = 0.05; 95% CI 0.01-0.24; corrected p < 0.0001) and ATID versus OT (OR = 1.22; 95% CI 0.45-3.18; corrected p = 1.00). Because genetic groups are strongly structured by geography, these comparisons were repeated after restriction to infections acquired in Europe and Central Asia, where type II accounted for 95.7% (859/898) of CT cases, 86.9% (146/168) of ATID cases, 73.7% (14/19) of OT cases and 42.9% (3/7) of ATIC cases. The differences between CT and the other categories persisted in this restricted analysis (CT versus ATID, corrected p = 0.0004; CT versus ATIC, corrected p = 0.0008; CT versus OT, corrected p = 0.009), indicating that they are not solely attributable to the geographical origin of infection.

Cases of no-CT were uncommon (n = 9) (S4 Table). Six were associated with type II strains, out of 875 type II strains recovered in the context of suspected or confirmed CT, whereas three were associated with type I strains, of which only five were identified in the entire series. Given the very small number of type I strains in this series, no statistical comparison was performed.

#### 3.5.1. Congenital toxoplasmosis

Among the 955 CT cases, clinical data were documented for 877 cases (91.8%). Most documented cases were infraclinical (645/877, 73.5%), whereas 58 (6.6%) presented with mild forms, 151 (17.2%) with severe forms and 23 (2.6%) with severe ocular toxoplasmosis, giving a combined proportion of severe or severe ocular forms of 19.8% (174/877) (S5 Table). Among cases with a documented trimester of maternal infection, 13.3% (123/927) occurred during the first trimester, 38.2% (354/927) during the second and 43.4% (402/927) during the third; a further 15 infections spanned the first and second trimesters, 24 the second and third, and 9 occurred before conception, in the periconceptional period, or in the context of maternal reactivation or reinfection.

The distribution of clinical forms among type II-associated CT cases broadly mirrored that of the overall CT cohort, with most cases being infraclinical (606/796 documented cases, 76.1%) and 134/796 (16.8%) presenting with severe or severe ocular forms. Severe and severe ocular forms type II-associated cases followed maternal infection in all three trimesters (67 after first-trimester, 41 after second-trimester and 12 after third-trimester infection), although the proportion of severe and severe ocular forms decreased sharply with gestational age at maternal infection, from 65.7% (67/102) in the first trimester to 13.6% (41/301) in the second and 3.6% (12/336) in the third (S6 Table). Type III strains showed a broadly similar clinical distribution, with most cases being infraclinical (16/18 documented cases, 88.9%) and 2/18 (11.1%) presenting with severe forms. In contrast, haplogroup 16 genotypes were predominantly associated with severe manifestations, with six of seven cases presenting with either severe CT (n = 4) or severe ocular CT (n = 2), and four of the six Africa 1-associated cases were also severe. The MCA for CT cases (Fig. 3) was performed on the cases for which genotypic, geographic, clinical, and gestational timing data were available. The MCA highlighted a proximity between infections acquired during the second and third trimesters and infraclinical or severe ocular forms. Conversely, infections occurring during the first trimester appeared to be more closely associated with severe forms. Regarding genetic groups, type II strains appeared to cluster closer to infraclinical, mild, and severe OT forms, whereas other genetic groups, such as haplogroup 16 or type III, appeared to occupy an intermediate position between these forms and severe forms.

**Fig. 3.**
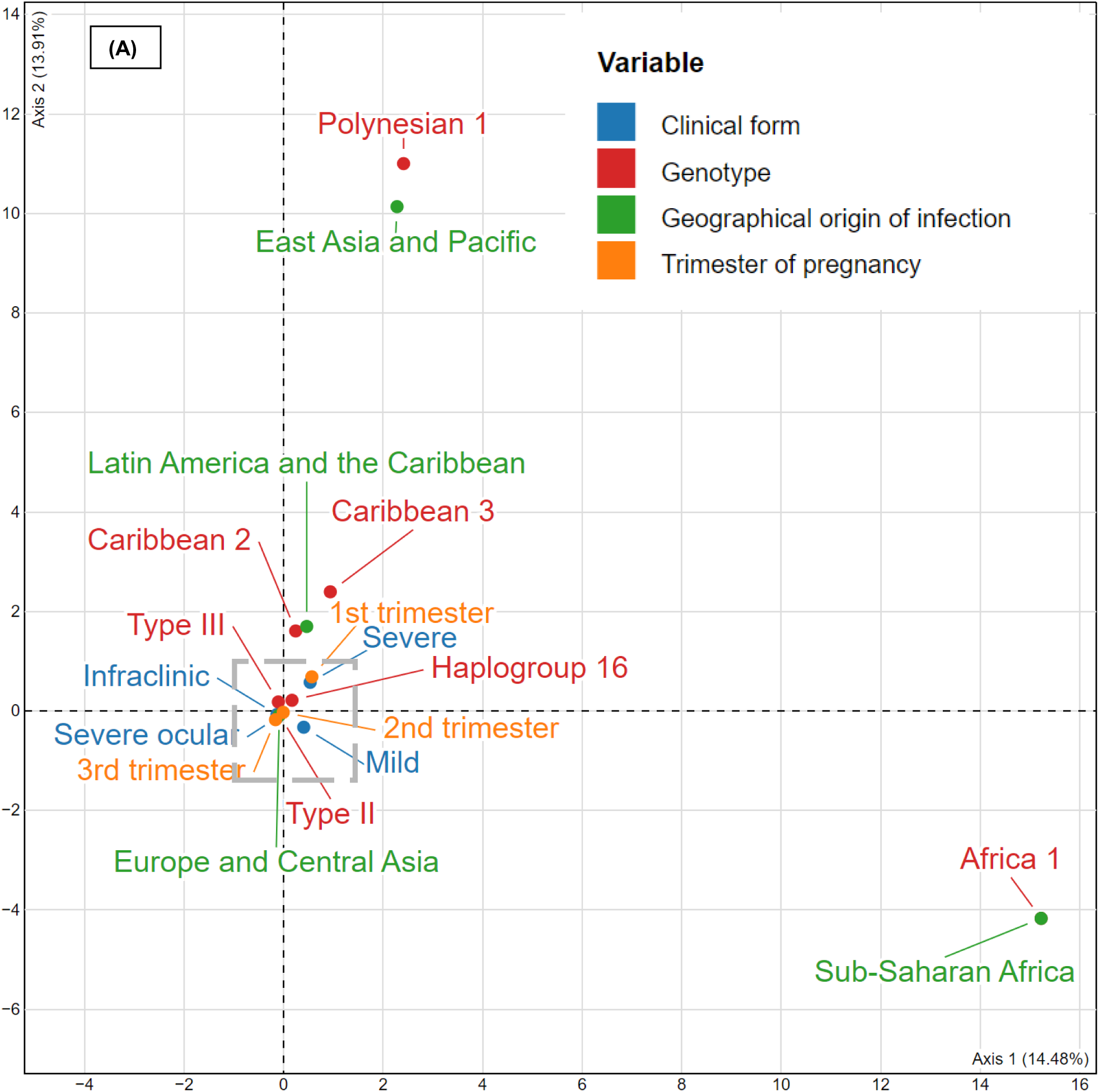
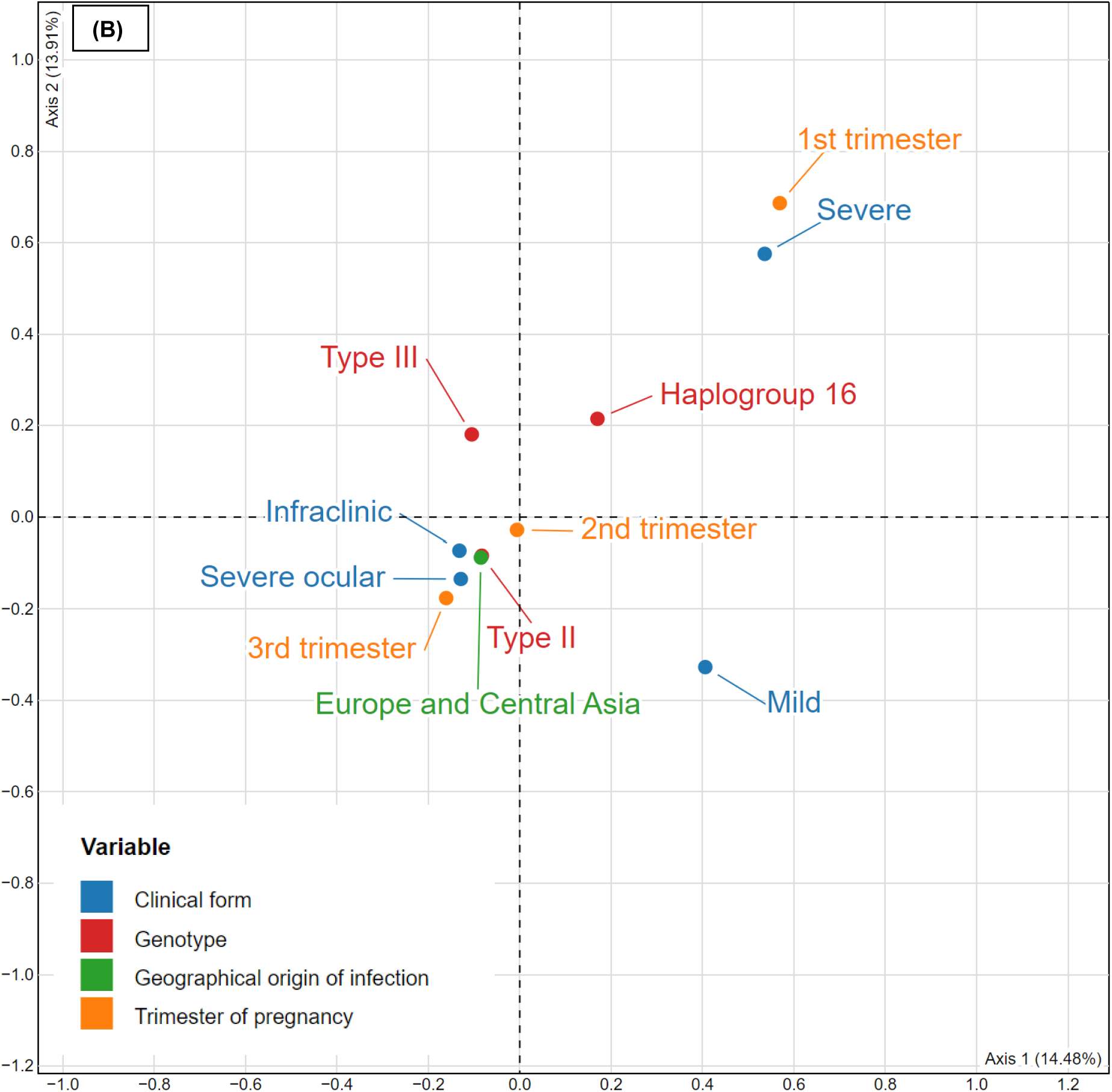
Multiple correspondence analysis (MCA) of congenital toxoplasmosis cases. Each point represents a category (modality) of the variables included in the analysis. The proximity between points reflects the tendency of these modalities to co-occur within the same cases, whereas distant points indicate weaker associations. Because complete disjunctive coding mechanically deflates the raw proportions of inertia, adjusted inertias are reported and the analysis is used for descriptive and hypothesis-generating purposes only. **(A)** Full view. **(B)** Enlarged view of the boxed region in panel A, showing the relative positions of the principal *Toxoplasma gondii* genetic groups and the clinical, geographical and gestational categories located in their immediate vicinity.

Nine CT cases followed maternal infection acquired before conception or in the periconceptional period, or occurred in the context of maternal reactivation. Six of these nine cases presented with severe forms, and seven were associated with type II strains. Although few in number, these observations indicate that congenital transmission and severe CT are not restricted to maternal infections acquired during pregnancy.

Type II, type III, and haplogroup 16 strains showed similar distributions across trimesters of maternal infection (S5 Table), so that the differences observed in the distribution of clinical forms between these groups are unlikely to be explained by gestational timing. Their geographical distributions differed, however: 98.8% of type II-associated CT cases were acquired in Europe and Central Asia, compared with 71.4% of type III-associated cases, so that the comparison between type II and type III strains remains confounded by the place of acquisition of infection. Comparative statistical analyses, using type II strains as the reference group and restricted to cases with documented clinical data, showed that haplogroup 16 strains were significantly more frequently associated with severe or severe ocular forms than type II strains (6/7 versus 134/796; OR = 29.5; 95% CI 3.53-1365; p = 0.0002, Bonferroni-corrected p = 0.0003). In contrast, no significant difference was observed between type III and type II strains (2/18 versus 134/796; OR = 0.62; 95% CI 0.07-2.68; p = 0.75). This comparison rests on only seven haplogroup 16 cases, with a very wide confidence interval, and should therefore be regarded as preliminary.

#### 3.5.2. Acquired toxoplasmosis in immunodeficient patients

Among the 238 ATID cases, disseminated (126/238, 52.9%) and isolated cerebral forms (99/238, 41.6%) predominated, whereas only 13 cases (5.5%) were paucisymptomatic or asymptomatic (S7 Table). HIV infection (111/238, 46.6%) and haematological malignancies or haematopoietic stem cell transplantation (82/238, 34.5%) were the main causes of immunodeficiency, followed by solid organ transplantation (28/238, 11.8%).

Isolated cerebral forms were far more frequent among patients infected with Africa 1 strains (20/25, 80.0%) than among those infected with type II (53/156, 34.0%) or type III strains (4/17, 23.5%). Conversely, disseminated forms predominated among type II-(90/156, 57.7%) and type III-associated cases (13/17, 76.5%). The distribution of immunodeficiency causes differed across these major genetic groups: haematological malignancies or stem cell transplantation predominated among type II-associated cases (45.5%), whereas HIV infection predominated among type III-(52.9%) and Africa 1-associated cases (80.0%). The MCA for ATID cases (Fig. 4) was performed on the cases for which genotypic, geographic, clinical, and immunodeficiency data were available. The MCA highlighted two main clusters. One cluster showed a relative proximity between type II strains, paucisymptomatic and disseminated forms, Europe and Central Asia location, haematological malignancies or haematopoietic stem cell transplantation, and SOT. The second cluster grouped type III and Africa 1, 2, and 4 strains with isolated cerebral forms and HIV infection, Sub-Saharan location and immunosuppressive or cytotoxic treatment. Comparative analyses showed no significant difference in the distribution of clinical forms between type II and type III strains (OR = 1.67; 95% CI 0.48-7.37; p = 0.59) (S7 Table). In contrast, isolated cerebral forms were significantly more frequent among patients infected with Africa 1 strains than among those infected with either type II strains (OR = 7.68; 95% CI 2.61-27.69; p < 0.0001) or type III strains (OR = 12.00; 95% CI 2.44-76.01; p = 0.0004). However, the distribution of causes of immunodeficiency differed between type II infections and most other genetic groups (S7 Table), suggesting that comparisons involving type II strains could be influenced by confounding related to immunodeficiency status: HIV infection was significantly more frequent among patients infected with Africa 1 strains than among those infected with type II strains (80.0% versus 32.7%; p < 0.0001). In contrast, no significant difference in the distribution of causes of immunodeficiency was observed between type III and Africa 1 strains (p = 0.08), although with 17 and 25 patients respectively this comparison cannot exclude differences of substantial magnitude. Nevertheless, Africa 1 strains remained significantly associated with isolated cerebral toxoplasmosis when compared with type III strains, despite broadly similar distributions of immunodeficiency causes between these two groups. This finding suggests that host immunodeficiency profiles alone may not fully account for the observed association, and supports a potential contribution of parasite genotype to the clinical presentation.

**Fig. 4.**
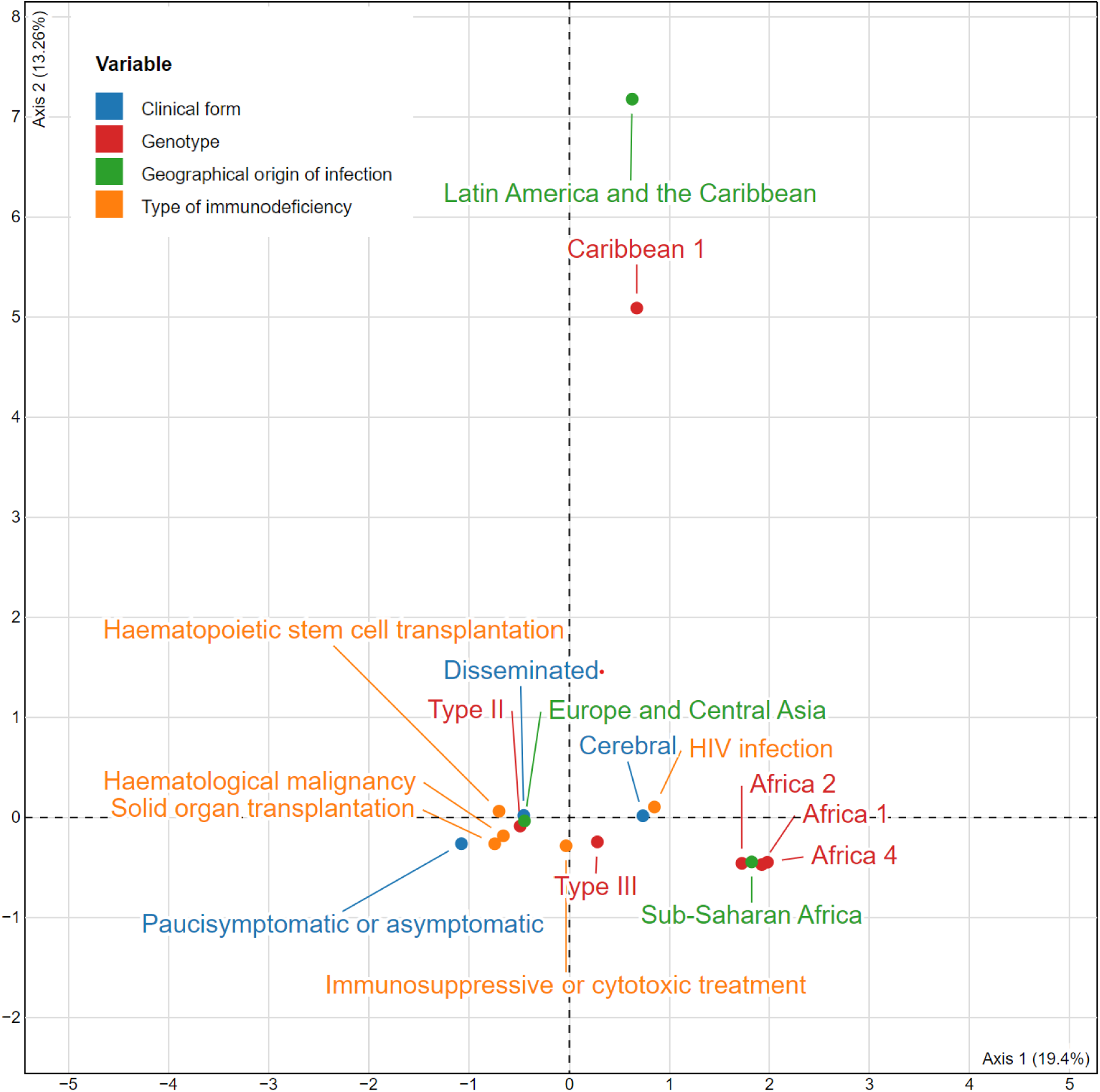
Multiple correspondence analysis (MCA) of acquired toxoplasmosis in immunodeficient patients. Each point represents a category (modality) of the variables included in the analysis, and proximity between points indicates a tendency for these modalities to co-occur within the same cases. **Adjusted inertias are reported and the analysis is used for descriptive and hypothesis-generating purposes only.**

#### 3.5.3. Acquired toxoplasmosis in immunocompetent patients

Among the 42 ATIC cases, 33 (78.6%) presented with severe clinical forms and 9 (21.4%) were paucisymptomatic (S8 Table). All 14 Amazonian-associated cases were severe; 12 had been acquired in French Guiana and 2 in Suriname. Twenty-three cases (54.8%) involved genotypes that could not be assigned to any recognized genetic group; 18 of these were severe and 19 had been acquired in Latin America and the Caribbean, a clinical and geographical distribution indistinguishable from that of the whole-genome-characterized Amazonian strains. Conversely, all three type II-associated cases and the single Africa 1-associated case were paucisymptomatic, and the single haplogroup 16-associated case was severe. Because the Amazonian group was defined on the basis of strains previously characterized by whole-genome sequencing, most of which originate from previously reported series of severe acquired toxoplasmosis in French Guiana, and because several ATIC strains from French Guiana shared identical 15-marker profiles within the same year and correspond to documented common-source outbreaks, no statistical comparison between genetic groups was performed for this clinical category and results are presented descriptively. One of these ATIC cases was the mother of a CT case included in the present series, the same strain having been recovered from the mother’s blood and from the fetal brain.

#### 3.5.4. Ocular toxoplasmosis

Among the 23 isolated OT cases, 8 lacked detailed clinical information or had no signs suggestive of severity, such as extensive inflammation (e.g., panuveitis), bilateral involvement, recurrence, or incomplete clinical improvement (S9 Table). These cases included 6 type II strains and 2 unclassified genotypes. In contrast, fifteen OT cases presented at least one clinical feature suggestive of severity. These cases included 11 immunocompetent patients (IC) and 4 immunodeficient patients (ID), and were associated with 8 type II strains, 2 Africa 1 strains, 2 unclassified genotypes, 1 Caribbean 1 strain, 1 haplogroup 16 strain, and 1 Mascarene 1 strain. Overall, 16 of the 23 genotyped OT cases occurred in immunocompetent patients, 5 in immunodeficient patients and 2 in patients whose immune status was not documented. In the 7 IC patients infected with type II strains, severe clinical presentations included bilateral granulomatous panuveitis, extensive retinitis, and persistent inflammation. These patients experienced recurrent episodes, sometimes over multiple years, or incomplete clinical improvement despite treatment. Because of the limited number of OT cases and the high diversity of genetic groups represented, no statistical analyses were performed for this clinical category.

## 4. Discussion

The nationwide French network of hospital parasitology diagnostic laboratories, officially constituted with the NRCT in 2006 for the epidemiological survey of congenital toxoplasmosis (CT), has collected 2,467 *T. gondii*-positive human clinical cases recorded between 1962 and 2023. The 1,269 *T. gondii* genotypes successfully characterized using 15 MS markers revealed a remarkably high genetic diversity, corresponding to 13 distinct genetic groups, including both clonal lineages and non-clonal populations, together with 79 genotypes that could not be assigned to any recognized group and that probably encompass a variety of rare lineages.

Several of these genetic groups have previously established correspondences with ToxoDB PCR-RFLP genotypes, considering that correspondences between microsatellite lineages and ToxoDB genotypes are not always one-to-one [37]. Type I, II, and III correspond to ToxoDB#10, #1-3, and #2, respectively. Africa 1 corresponds to ToxoDB#6, Africa 4 to ToxoDB#20 and #137, and haplogroup 16 to multiple genotypes including ToxoDB#15, #17, and #202. Caribbean lineages correspond mainly to ToxoDB#13 (Caribbean 1), #12 and #31 (Caribbean 2), and #25 and #141 (Caribbean 3), whereas Amazonian strains encompass multiple genotypes including ToxoDB#60, #95, #97, #98, #193, and #194. The Mascarene 1 lineage corresponds to ToxoDB#99 [38].

This high level of genetic diversity can be explained by several factors. First, the nationwide French network of hospital parasitology diagnostic laboratories collect clinical samples not only mainland France, but also French overseas territories and collectivities located in South America (French Guiana), the Caribbean archipelago (Guadeloupe and Martinique) the Indian Ocean (Réunion Island) and Oceania (French Polynesia and New Caledonia). The numerous genotypes identified in residents of French overseas territories and collectivities reflected the local genetic diversity of these areas, including Caribbean 1, 2, and 3 lineages in the Caribbean islands and French Guiana, Amazonian strains in French Guiana, the Mascarene 1 lineage in Réunion Island, and the Polynesian 1 lineage in French Polynesia in Oceania. Second, clinical samples collected in mainland France also included samples from immigrants and travelers receiving care at French hospitals. Because a substantial proportion of immigrants living in France originate from Africa [39], African lineages, mainly Africa 1 but also Africa 2 and Africa 4, were well represented among the *T. gondii* genotypes characterized through the activities of the French network. Notably, most of these lineages were described for the first time through the activities of the French nationwide network [29,37,40]. Third, clinical samples collected in mainland France also included residents with no history of travel but with confirmed or suspected exposure to imported food products such as horsemeat. Notably, the association between toxoplasmosis and consumption of imported horse meat has been previously documented through the NRCT activities [8,41].

This particularly high genetic diversity observed in human samples contrasts markedly with that reported in domestic and wild animals in mainland France, in which *T. gondii* strains are overwhelmingly represented by the type II lineage, with only a single type III strain reported to date [42–45]. However, among human cases with Europe and Central Asia as the reported place of infection, genetic diversity was substantially lower and more closely resembled that documented in animal reservoirs, with type II accounting for 93.3% of cases.

Within each clinical category (CT, ATID, ATIC, and OT), the number of collected and successfully genotyped *T. gondii-positive* clinical samples and isolates, as well as the diversity of *T. gondii* genotypes identified, varied substantially. Cases of CT accounted for the largest proportion of genotyped samples. In France, public health policies recommend systematic screening of pregnant women at the beginning of pregnancy, followed by regular serological monitoring throughout gestation in seronegative women, even in the absence of clinical symptoms [46,11]. This emphasis on CT surveillance is reflected in the activities of the NRCT, which was established primarily to conduct epidemiological surveillance of CT infections. Most *T. gondii* genotypes identified in cases of CT belonged to the type II lineage, probably reflecting local *T. gondii* diversity in mainland France. However, a wide range of other genetic groups was also detected, reflecting infections acquired in overseas territories, among travelers and immigrants, cases associated with the consumption of imported horse meat, and infections for which no clear source of exposure could be identified. Cases of ATID accounted for a substantial proportion of genotyped samples. Although type II remained the predominant lineage in this clinical category, the Africa 1, Africa 2, Africa 4, and type III lineages were also well represented. These genotypes were frequently identified in patients originating from Africa who were living in France and presented with reactivation of a latent *T. gondii* infection. This pattern is consistent with the circulation of these lineages in several African regions and with the higher burden of HIV infection reported in some of these settings. Cases of ATIC were less frequent, reflecting the fact that *T. gondii* infection is most often asymptomatic or paucisymptomatic in immunocompetent individuals in mainland France [47]. Most genotyped strains from this category were acquired in French Guiana and belonged either to the whole-genome-characterized Amazonian population or to unassignable genotypes sharing the same geographical origin; this population is associated with exposure to the wild environment of French Guiana and is known to cause severe disseminated toxoplasmosis in immunocompetent hosts [48,49]. Cases of isolated OT were also uncommon. This likely reflects both the limited success of genotyping directly from ocular samples and the near impossibility of isolating viable strains from these specimens. Although type II accounted for more than half of the genotyped OT cases (60.9%), its proportion was markedly lower than that observed in CT (91.0%). A variety of other genetic groups were also represented. This pattern may reflect differences in the higher propensity of certain *T. gondii* genetic groups to cause OT compared with type II strains, although the limited number of OT cases precluded meaningful statistical comparisons.

The results of the present study, based on the largest collection of human *T. gondii* genotypes assembled to date, indicate that a given *T. gondii* genetic group can be associated with a wide spectrum of clinical presentations. Nevertheless, significant differences in the distribution of clinical manifestations and in the frequency of severe forms were observed between certain genetic groups.

Among CT cases, type II strains were predominantly associated with infraclinical or mild forms. However, a substantial proportion of type II-associated CT cases presented with severe manifestations (16.8% of cases with documented clinical data). These included infections acquired before conception or during early pregnancy, when clinical severity is generally considered to be driven primarily by gestational timing rather than parasite genotype, but also infections acquired during later stages of pregnancy, where parasite genetic factors may play a more important role in determining clinical outcome [50,51]. Nine CTcases followed maternal infection acquired before conception or in the periconceptional period, or occurred after maternal reactivation, and six of these were severe; although based on small numbers, this observation is of interest given that the previously reported haplogroup 16 case of severe CT also followed maternal reinfection during pregnancy [8]. Compared with type II strains, haplogroup 16 strains were significantly associated with severe CT, whereas no such association was observed for type III strains. This association rests on seven cases only, with very wide confidence intervals, and require confirmation in larger series. Haplogroup 16 is a poorly described genetic group; it was first defined through whole-genome phylogenetic analysis of 62 *T. gondii* genomes from diverse geographic origins, in which it formed a well-supported, genetically distinct branch [32]. Among the strains assigned to this haplogroup were three from patients investigated through the French NRCT: the previously reported IPP-URB strain (TgH20005), responsible for a severe CT case following maternal reinfection during pregnancy associated with imported horsemeat consumption [52], and two additional strains (TgH26044 and TgH21016) isolated from patients with no history of travel or reported consumption of imported horse meat. Additional haplogroup 16 strains have repeatedly been identified through NRCT microsatellite genotyping, all carrying the characteristic TUB2 allele 287. Although its geographical origin remains elusive, several observations nevertheless point towards a South American origin for this genetic group, even though all ten haplogroup 16 infections of the present series were acquired in Europe. CASTELLS, the only strain in the original whole-genome dataset with a documented geographic origin outside the French NRCT, was isolated from a sheep in Uruguay [53]. Moreover, although haplogroup 16 forms a distinct phylogenetic branch, whole-genome analyses indicate genetic proximity to South American genomes [31]. These observations are of particular interest given the high burden and severity of toxoplasmosis reported in South America [6]. Although parasite genetic diversity is thought to contribute to this pattern, the genetic groups involved remain poorly characterized [54]. Our findings suggest that haplogroup 16 may represent one such group and warrant further investigation, particularly in the context of CT.

The associations discussed so far concern the severity of CT once the parasite has reached the fetus. Whether genetic groups also differ in their ability to reach the fetus at all is a distinct question. In the present study, all nine cases in which CT was initially suspected but mother-to-child transmission was ultimately excluded correspond to a strain isolated from the placenta of a child who was not infected (no-CT), and their genotype distribution is unbalanced: six arose among the 875 type II strains recovered from suspected or confirmed CT, whereas three arose among the five type I strains of the entire collection. Taken together, these five strains are too few to support any inference on the transmissibility of type I strains to the fetus, or to argue that they cross the placenta less efficiently than type II strains. Elsewhere, in a previous study, the rate of CT has been reported higher in Martinique than in mainland France, suggesting that certain *T. gondii* lineages from Caribbean islands (i.e., Caribbean 1, 2, and 3) could be more efficiently transmitted to the fetus than type II strains [55].

In cases of acquired toxoplasmosis in immunodeficientpatients (ATID), compared with type II strains, which were more frequently associated with disseminated forms of toxoplasmosis, Africa 1 strains were more frequently associated with isolated cerebral forms, whereas type III strains showed a distribution of clinical forms similar to that observed for type II strains. A previous study based on a smaller series of ATID cases from the French NRCT and the associated nationwide network attributed the high frequency of cerebral forms among patients infected with Africa 1 strains to the predominance of HIV infection in this group [29], as HIV-positive patients are more likely to present with cerebral toxoplasmosis than patients with other causes of immunodeficiency. This hypothesis was supported by the fact that HIV infection was significantly more frequent among patients infected with Africa 1 strains (80.0%) than among those infected with type II strains (32.7%). The association between cerebral forms and HIV infection was subsequently supported by another study [53]. However, the present study provides additional insight. Although HIV infection was also the predominant cause of immunodeficiency among patients infected with type III strains (52.9%), no significant difference in the distribution of causes of immunodeficiency was observed between Africa 1 and type III infections, although with 25 and 17 patients respectively this comparison cannot exclude differences of substantial magnitude. Despite these comparable immunodeficiency profiles, isolated cerebral forms remained significantly more frequent among patients infected with Africa 1 strains than among those infected with type III strains. This observation suggests that differences in immunodeficiency status alone are unlikely to fully explain the clinical differences observed between these two genetic groups. Taken together, these findings support an association between Africa 1 strains and isolated cerebral forms in immunodeficient patients and suggest that *T. gondii* genotype may play a role in determining the clinical presentation of toxoplasmosis even in the context of immunodeficiency. Nevertheless, this interpretation should be considered with caution, given the relatively small sample sizes for these lineages and the potential influence of other factors that could not be assessed in the present study, including host genetic background or ethnicity, which have previously been implicated in the patterns observed for OT and ATIC cases [10,18].

In cases of acquired toxoplasmosis in immunocompetent patients (ATIC), type II strains were uncommon and were identified only in paucisymptomatic cases. In contrast, most cases were associated with strains acquired in French Guiana, which belong to a *T. gondii* population with several unique characteristics. This population is almost exclusively associated with the wild environment of French Guiana and displays exceptionally high genetic diversity. Despite this diversity, WGS studies have demonstrated that Amazonian strains constitute a well-defined discrete genetic group [31,32]. The present study shows, however, that this group cannot be delimited with 15 microsatellite markers, and a majority of the ATIC cases of this series involved genotypes that could not be assigned to any recognized group but whose geographical origin was indistinguishable from that of whole-genome-characterized Amazonian strains. Taken together, these strains were almost invariably associated with severe disseminated toxoplasmosis, in keeping with the case reports and case series coordinated by members of the French network that have documented the exceptional pathogenicity of this population, which can be life-threatening if left untreated in immunocompetent individuals exposed to the Amazonian rainforest environment [48,49]. In addition, haplogroup 16 was also associated with one severe disease case in an immunocompetent patient. While haplogroup 16 was more strongly associated with severe CT in the present study, this observation suggests that its pathogenic potential may not be restricted to CT. Of note, the previously reported case of severe CT caused by a haplogroup 16 strain occurred following maternal reinfection during pregnancy in a woman who had been infected before conception and experienced a paucisymptomatic clinical presentation [52]. Although based on very limited numbers, these observations raise the possibility that haplogroup 16 may exhibit an intermediate level of pathogenicity between type II and Amazonian strains. The single Africa 1 strain identified among ATIC cases was associated with a paucisymptomatic presentation. Interestingly, although the Africa 1 lineage is primarily associated with ATID cases, suggesting a lower apparent pathogenicity in immunocompetent hosts, a small case series of ATIC associated with Africa 1 strains has previously been published by the French nationwide network in immunocompetent immigrants from Africa and travelers returning from Africa [56]. Of note, one immunocompetent patient infected with an Africa 1 strain from this study had severe disseminated toxoplasmosis; however, that infection was associated with an incomplete genotype and could therefore not be included in the present analyses.

In patients with OT, most ocular samples could not be successfully genotyped, likely because of the low *T. gondii* DNA burden typically present in ocular specimens, in particular aqueous humor; only 7.1% of the 325 OT cases of the series yielded a complete genotype. This previously observed limitation [57,58] introduces a major source of bias when investigating associations between *T. gondii* genotype and clinical presentation. Among successfully genotyped cases, type II strains accounted for more than half of the genotypes identified, but a substantial proportion of OT cases also encompassed a broad range of other genetic groups (Africa 1, Caribbean 1, haplogroup 16, Mascarene 1 and unclassified genotypes), collectively accounting for a substantial proportion of cases, although each group was represented by only a small number of cases. Although the limited number of successfully genotyped samples precludes firm conclusions, this observation may suggest a greater propensity of certain non-type II genotypes to proliferate in ocular tissues. Conversely, several OT cases associated with type II strains in both immunocompetent and immunodeficient patients presented features commonly associated with severe disease, including bilateral involvement, extensive inflammation, recurrent episodes, or incomplete clinical improvement despite treatment. This observation is consistent with previous findings from the French nationwide network [57], which demonstrated that type II strains can cause severe forms of OT. It should be noted, however, that most cases classified as severe met the criterion of recurrent or reactivated disease (14 of 15). Because recurrence can only be observed in patients followed long enough, and follow-up duration was not documented in this dataset, patients seen on a single occasion cannot be identified as severe on this criterion. The relative frequency of severe OT according to *T. gondii* genetic group therefore remains largely unknown. Addressing this question will require larger datasets and improved genotyping success rates, while also accounting for the multiple other factors that may influence disease severity.

The present study has several limitations related to its retrospective design and to the fact that the dataset was not initially established for genotype-clinic association analyses. Consequently, data collection was not fully standardized over time, resulting in missing data for several variables. In addition, analyses were performed separately for each clinical category, leading to marked imbalances in sample sizes across categories and genetic groups. This limited the statistical power of certain analyses and contributed to wide confidence intervals and large odds ratios for some comparisons. Furthermore, due to the nature of the dataset, which was derived from cases analyzed by the French NRCT, some genetic groups of *T. gondii* that are rare or exotic in France were represented by only a small number of cases within each clinical category. This limited representation reduced the statistical power of the analyses and precluded robust assessment of whether certain rare genetic groups are associated with increased disease severity. Missing data also prevented the implementation of multivariable analyses and may have introduced residual confounding. Although potential confounding factors were partially explored through MCA and subsequent pairwise statistical comparisons, the influence of unmeasured variables cannot be excluded. An additional limitation relates to the strong correlation observed between certain *T. gondii* genetic groups and the geographic origin of infection. Several lineages showed highly structured geographic distributions, including Amazonian strains in French Guiana, Africa 1 strains in sub-Saharan Africa, and Caribbean lineages in the French Caribbean territories. Consequently, the respective contributions of parasite genotype, geographic origin, environmental exposure, host genetic background, and other region-specific epidemiological factors could not be fully disentangled. Although geographic origin was incorporated into the MCA, the absence of multivariable models prevented formal adjustment for these potentially confounding factors; the comparisons restricted to infections acquired in Europe and Central Asia provide only a partial answer to this question. Similarly, some host-related variables were unevenly distributed across genetic groups. For example, HIV infection predominated among patients infected with Africa 1 strains, whereas haematological malignancies were more frequent among patients infected with type II strains. Although these factors were considered in the exploratory analyses and further investigated through targeted pairwise comparisons, residual confounding cannot be excluded. Moreover, because the multiple correspondence analyses and the confirmatory tests were performed on the same dataset, the corresponding p-values should be interpreted as exploratory. Another important limitation relates to genotyping success, which varied substantially according to the type of clinical sample analyzed. While genotyping success was high for isolated strains, it was considerably lower for primary clinical specimens and particularly limited for ocular samples. This differential genotyping success may have introduced selection bias if certain *T. gondii* genotypes are associated with higher parasite burdens or are more readily amplified from clinical specimens. This recurring limitation in *T. gondii* molecular epidemiology highlights the need to develop improved parasite enrichment and purification methods for low-parasite-load clinical samples. Finally, it should be noted that the French NRCT is a surveillance network rather than a population-based cohort. As a result, uncommon, severe, or clinically atypical cases are more likely to be investigated and genotyped than mild infections. Consistent with this, complete genotyping was more frequent for severe or clinically overt presentations in all clinical categories except ocular toxoplasmosis, so that the clinical spectrum associated with each genetic group may be shifted towards its more symptomatic end. This potential recruitment bias may be particularly relevant for ATIC patients, for which severe forms are more likely to be referred to specialized centers and included in the NRCT database.

## 5. Conclusion

In conclusion, this study, based on the largest collection of human *T. gondii* genotypes assembled to date, highlights the remarkable diversity of parasite strains infecting humans receiving care in France, and illustrates the value of long-term national surveillance networks for investigating the epidemiology and clinical impact of toxoplasmosis. By combining standardized microsatellite-based genotyping with clinical and epidemiological information collected over more than four decades, the French network of hospital parasitology diagnostic laboratories provided a unique opportunity to investigate the relationships between *T. gondii* genotypes and the clinical manifestations of different forms of human toxoplasmosis in patients infected in France or in other regions of the world. Our findings indicate that although a given *T. gondii* genetic group can be associated with a broad spectrum of clinical manifestations, certain genetic groups display preferential associations with specific disease presentations. Type II strains, while predominantly associated with infraclinical or mild CT, were also responsible for severe CT following maternal infection in all three trimesters of pregnancy, as well as for severe ocular disease. Type I strains were remarkably rare in this collection and showed no signs of increased clinical severity compared to type II. Amazonian strains were identified almost exclusively in immunocompetent patients with severe acquired toxoplasmosis, in keeping with previous reports of the exceptional pathogenicity of this genetic group. Haplogroup 16 emerged as a genetic group associated with severe CT. In immunodeficient patients, Africa 1 strains showed a particular association with isolated cerebral forms compared to type II and type III strains. Taken together, these findings suggest a role for parasite genotype in shaping the clinical expression of toxoplasmosis, even in the presence of strong host-related determinants such as immunodeficiency or gestational timing of infection. Finally, our results emphasize the need to expand genetic surveillance internationally and pursue whole-genome investigations across diverse geographic regions to better characterize the global diversity of *T. gondii* and understand the biological determinants underlying the pathogenicity of its different genetic groups.

## Data Availability

All data produced in the present work are contained in the manuscript

## Acknowledgments

We would like to thank all public health professionals involved in data collection and all study participants. We thank the French Agence Nationale de la Recherche (ToxoPath ANR-25-CE35-7959) for providing research funds to Lokman Galal.

## Notes

### Competing Interest Statement

The authors have declared no competing interest.

### Author Declarations

The analysis of biological samples obtained through standard clinical practices was classified as non-interventional research in accordance with Article L1221-1.1 of the French Public Health Code, requiring only the absence of patient objection at the time of sample collection (as per Article L1211-2 of the Public Health Code). At participating centers, mothers and patients were informed about the anonymized reporting of CT cases, the collection of clinical and epidemiological data, and the study of T. gondii isolates from different clinical forms of toxoplasmosis. Mothers and patients had the right to refuse participation in this survey. All collected data were anonymized before samples were sent to the Limoges laboratory. Anonymized clinical and epidemiological data associated with T. gondii strains collected through the French NRCT were reported by the participating specialized laboratories via a dedicated online data-entry system (http://cnrtoxoplasmose.chu-reims.fr), using the specifically developed ToxoSurv software (EpiConcept). The software and associated data collection procedures were approved by the French National Data Protection Authority (Commission Nationale de l Informatique et des Libertes, CNIL; approval reference DR-2016-186).

## References

1. Toxoplasmosis Montoya, JG, et al. The Lancet, Volume 363, Issue 9425, 1965–1976.

2. Sengupta PP, Jacob SS, Suresh KP, Rajamani S, Maharana SM. Exploring global trends in human toxoplasmosis seroprevalence by meta-analysis. Experimental Parasitology. 2025;275: 108971.

3. Hill D, Dubey JP. Toxoplasma gondii: transmission, diagnosis and prevention. Clinical Microbiology and Infection. 2002;8: 634–640. doi:10.1046/j.1469-0691.2002.00485.x

4. Howe DK, Sibley LD. Toxoplasma gondii Comprises Three Clonal Lineages: Correlation of Parasite Genotype with Human Disease. Journal of Infectious Diseases. 1995;172: 1561–1566. doi:10.1093/infdis/172.6.1561

5. Owen MR, Trees AJ. Genotyping of Toxoplasma gondii Associated with Abortion in Sheep. The Journal of Parasitology. 1999;85: 382. doi:10.2307/3285654

6. Gilbert RE, Freeman K, Lago EG, Bahia-Oliveira LMG, Tan HK, Wallon M, et al. Ocular Sequelae of Congenital Toxoplasmosis in Brazil Compared with Europe. Campos MAS, editor. PLoS Negl Trop Dis. 2008;2: e277. doi:10.1371/journal.pntd.0000277

7. De Salvador-Guillouët F, Ajzenberg D, Chaillou-Opitz S, Saint-Paul M-C, Dunais B, Dellamonica P, et al. Severe pneumonia during primary infection with an atypical strain of Toxoplasma gondii in an immunocompetent young man. Journal of Infection. 2006;53: e47–e50. doi:10.1016/j.jinf.2005.10.026

8. Elbez-Rubinstein A, Ajzenberg D, Dardé M, Cohen R, Dumètre A, Yera H, et al. Congenital Toxoplasmosis and Reinfection during Pregnancy: Case Report, Strain Characterization, Experimental Model of Reinfection, and Review. J INFECT DIS. 2009;199: 280–285. doi:10.1086/595793

9. de-la-Torre A, Mejía-Salgado G, Eraghi AT, Pleyer U. Age and ocular toxoplasmosis: a narrative review. FEMS Microbes. 2025;6: xtaf002. doi:10.1093/femsmc/xtaf002

10. Rudzinski M, Khoury M, Couto C, Ajzenberg D. Reactivation of Ocular Toxoplasmosis in Non-Hispanic Persons, Misiones Province, Argentina. Emerg Infect Dis. 2016;22: 912–913. doi:10.3201/eid2205.150025

11. Mandelbrot L, Kieffer F, Wallon M, Winer N, Massardier J, Picone O, et al. Toxoplasmose pendant la grossesse : proposition actuelle de prise en charge pratique. Gynécologie Obstétrique Fertilité & Sénologie. 2021;49: 782–791. doi:10.1016/j.gofs.2021.03.003

12. Darde ML, Bouteille B, Pestre-Alexandre M. Isoenzymic characterization of seven strains of Toxoplasma gondii by isoelectrofocusing in polyacrylamide gels. The American journal of tropical medicine and hygiene. 1988;39: 551–558.

13. Dardé ML, Bouteille B, Pestre-Alexandre M. Isoenzyme analysis of 35 Toxoplasma gondii isolates and the biological and epidemiological implications. The Journal of parasitology. 1992; 786–794.

14. Ajzenberg D, Bañuls A-L, Tibayrenc M, Dardé ML. Microsatellite analysis of Toxoplasma gondii shows considerable polymorphism structured into two main clonal groups. International Journal for Parasitology. 2002;32: 27–38. doi:10.1016/S0020-7519(01)00301-0

15. Ajzenberg D, Collinet F, Mercier A, Vignoles P, Dardé M-L. Genotyping of Toxoplasma gondii Isolates with 15 Microsatellite Markers in a Single Multiplex PCR Assay. J Clin Microbiol. 2010;48: 4641–4645. doi:10.1128/JCM.01152-10

16. Su C, Shwab EK, Zhou P, Zhu XQ, Dubey JP. Moving towards an integrated approach to molecular detection and identification of Toxoplasma gondii. Parasitology. 2010;137: 1–11. doi:10.1017/S0031182009991065

17. Shwab EK, Jiang T, Pena HFJ, Gennari SM, Dubey JP, Su C. The ROP18 and ROP5 gene allele types are highly predictive of virulence in mice across globally distributed strains of Toxoplasma gondii. Int J Parasitol. 2016;46: 141–146. doi:10.1016/j.ijpara.2015.10.005

18. Vidal N, Faucher J-F, Blaizot R, Pommier de SAnti V, Valdes A, Tabi R, et al. Toxoplasmosis acquired in French Guiana in immunocompetent adults: case series from 2002 to 2019, with a focus on outpatients. Open Forum Infectious Diseases. Oxford University Press US; 2026. p. ofag087.

19. Hosseini SA, Amouei A, Sharif M, Sarvi Sh, Galal L, Javidnia J, et al. Human toxoplasmosis: a systematic review for genetic diversity of *Toxoplasma gondii* in clinical samples. Epidemiol Infect. 2019;147: e36. doi:10.1017/S0950268818002947

20. Ajzenberg D, Cogné N, Paris L, Bessières M, Thulliez P, Filisetti D, et al. Genotype of 86 *Toxoplasma gondii* Isolates Associated with Human Congenital Toxoplasmosis, and Correlation with Clinical Findings. J INFECT DIS. 2002;186: 684–689. doi:10.1086/342663

21. Khan A, Jordan C, Muccioli C, Vallochi AL, Rizzo LV, Belfort R, et al. Genetic Divergence of *Toxoplasma gondii* Strains Associated with Ocular Toxoplasmosis, Brazil. Emerg Infect Dis. 2006;12: 942–949. doi:10.3201/eid1206.060025

22. Ajzenberg D. Type I Strains in Human Toxoplasmosis: Myth Or Reality? Future Microbiol. 2010;5: 841–843. doi:10.2217/fmb.10.55

23. Boughattas S, Ben-Abdallah R, Siala E, Souissi O, Aoun K, Bouratbine A. Direct Genotypic Characterization of Toxoplasma gondii Strains Associated with Congenital Toxoplasmosis in Tunisia (North Africa). The American Society of Tropical Medicine and Hygiene. 2010;82: 1041–1046. doi:10.4269/ajtmh.2010.09-0691

24. Vilares A, Gargaté MJ, Ferreira I, Martins S, Gomes JP. Molecular and virulence characterization of Toxoplasma gondii strains isolated from humans in Portugal. Parasitol Res. 2017;116: 979–985. doi:10.1007/s00436-017-5374-5

25. Lachkhem A, Galal L, Lahmar I, Passebosc K, Riahi H, Plault N, et al. First isolation and genotyping of Toxoplasma gondii strains from domestic animals in Tunisia. Sci Rep. 2021;11: 9328. doi:10.1038/s41598-021-88751-1

26. Villena I, Ancelle T, Delmas C, Garcia P, Brézin AP, Thulliez P, et al. Congenital toxoplasmosis in France in 2007: first results from a national surveillance system. Eurosurveillance. 2010;15. doi:10.2807/ese.15.25.19600-en

27. Lebech M, Joynson DHM, Seitz HM, Thulliez P, Gilbert RE, Dutton GN, et al. Classification system and case definitions of Toxoplasma gondii infection in immunocompetent pregnant women and their congenitally infected offspring. European Journal of Clinical Microbiology and Infectious Diseases. 1996;15: 799–805.

28. Ajzenberg D, Dumètre A, Dardé M-L. Multiplex PCR for Typing Strains of *Toxoplasma gondii*. J Clin Microbiol. 2005;43: 1940–1943. doi:10.1128/JCM.43.4.1940-1943.2005

29. Ajzenberg D, Yera H, Marty P, Paris L, Dalle F, Menotti J, et al. Genotype of 88 *Toxoplasma gondii* Isolates Associated with Toxoplasmosis in Immunocompromised Patients and Correlation with Clinical Findings. J INFECT DIS. 2009;199: 1155–1167. doi:10.1086/597477

30. Joeres M, Cardron G, Passebosc-Faure K, Plault N, Fernandez-Escobar M, Hamilton CM, et al. A ring trial to harmonize Toxoplasma gondii microsatellite typing: comparative analysis of results and recommendations for optimization. European Journal of Clinical Microbiology & Infectious Diseases. 2023;42: 803–818.

31. Galal L, Ariey F, Gouilh MA, Dardé M-L, Hamidović A, Letourneur F, et al. A unique Toxoplasma gondii haplotype accompanied the global expansion of cats. Nature Communications. 2022;13: 5778. doi:10.1038/s41467-022-33556-7

32. Lorenzi H, Khan A, Behnke MS, Namasivayam S, Swapna LS, Hadjithomas M, et al. Local admixture of amplified and diversified secreted pathogenesis determinants shapes mosaic Toxoplasma gondii genomes. Nat Commun. 2016;7: 10147. doi:10.1038/ncomms10147

33. Ayele D, Zewotir T, Mwambi H. Multiple correspondence analysis as a tool for analysis of large health surveys in African settings. Afr H Sci. 2015;14: 1036. doi:10.4314/ahs.v14i4.35

34. Costa PS, Santos NC, Cunha P, Cotter J, Sousa N. The Use of Multiple Correspondence Analysis to Explore Associations between Categories of Qualitative Variables in Healthy Ageing. Journal of Aging Research. 2013;2013: 1–12. doi:10.1155/2013/302163

35. Greenacre M, Blasius J, editors. Multiple Correspondence Analysis and Related Methods. New York: Chapman and Hall/CRC; 2006. doi:10.1201/9781420011319

36. Sourial N, Wolfson C, Zhu B, Quail J, Fletcher J, Karunananthan S, et al. Correspondence analysis is a useful tool to uncover the relationships among categorical variables. Journal of Clinical Epidemiology. 2010;63: 638–646. doi:10.1016/j.jclinepi.2009.08.008

37. Galal L, Hamidović A, Dardé ML, Mercier M. Diversity of Toxoplasma gondii strains at the global level and its determinants. Food and Waterborne Parasitology. 2019;15: e00052.

38. Su C, Khan A, Zhou P, Majumdar D, Ajzenberg D, Dardé M-L, et al. Globally diverse Toxoplasma gondii isolates comprise six major clades originating from a small number of distinct ancestral lineages. Proc Natl Acad Sci USA. 2012;109: 5844–5849. doi:10.1073/pnas.1203190109

39. Rouhban O. En 2023, 3, 5 millions d’immigrés nés en Afrique vivent en Franc. L’Institut national de la statistique et des etudes economiques. 2024.

40. Yera H, Ajzenberg D, Lesle F, Eyrolle-Guignot D, Besnard M, Baud A, et al. New description of Toxoplasma gondii genotypes from French Polynesia. Acta Tropica. 2014. pp. 10–12. doi:10.1016/j.actatropica.2014.02.011

41. Pomares C, Ajzenberg D, Bornard L, Bernardin G, Hasseine L, Darde M-L, et al. Toxoplasmosis and horse meat, France. Emerging Infect Dis. 2011;17: 1327–1328. doi:10.3201/eid1707.101642

42. Richomme C, Aubert D, Gilot-Fromont E, Ajzenberg D, Mercier A, Ducrot C, et al. Genetic characterization of Toxoplasma gondii from wild boar (Sus scrofa) in France. Vet Parasitol. 2009;164: 296–300. doi:10.1016/j.vetpar.2009.06.014

43. Aubert D, Ajzenberg D, Richomme C, Gilot-Fromont E, Terrier ME, de Gevigney C, et al. Molecular and biological characteristics of Toxoplasma gondii isolates from wildlife in France. Vet Parasitol. 2010;171: 346–349. doi:10.1016/j.vetpar.2010.03.033

44. Halos L, Thébault A, Aubert D, Thomas M, Perret C, Geers R, et al. An innovative survey underlining the significant level of contamination by Toxoplasma gondii of ovine meat consumed in France. Int J Parasitol. 2010;40: 193–200. doi:10.1016/j.ijpara.2009.06.009

45. Djokic V, Blaga R, Aubert D, Durand B, Perret C, Geers R, et al. Toxoplasma gondii infection in pork produced in France. Parasitology. 2016;143: 557–567. doi:10.1017/S0031182015001870

46. Peyron F, L’ollivier C, Mandelbrot L, Wallon M, Piarroux R, Kieffer F, et al. Maternal and congenital toxoplasmosis: diagnosis and treatment recommendations of a French multidisciplinary working group. Pathogens. 2019;8: 24.

47. Robert-Gangneux F, Dardé M-L. Epidemiology of and Diagnostic Strategies for Toxoplasmosis. Clin Microbiol Rev. 2012;25: 264–296. doi:10.1128/CMR.05013-11

48. Carme B, Bissuel F, Ajzenberg D, Bouyne R, Aznar C, Demar M, et al. Severe acquired toxoplasmosis in immunocompetent adult patients in French Guiana. J Clin Microbiol. 2002;40: 4037–4044.

49. Demar M, Hommel D, Djossou F, Peneau C, Boukhari R, Louvel D, et al. Acute toxoplasmoses in immunocompetent patients hospitalized in an intensive care unit in French Guiana. Clin Microbiol Infect. 2012;18: E221–231. doi:10.1111/j.1469-0691.2011.03648.x

50. Dunn D, Wallon M, Peyron F, Petersen E, Peckham C, Gilbert R. Mother-to-child transmission of toxoplasmosis: risk estimates for clinical counselling. The Lancet. 1999;353: 1829–1833.

51. Thiébaut R, Leproust S, Chêne G, Gilbert R. Effectiveness of prenatal treatment for congenital toxoplasmosis: a meta-analysis of individual patients’ data. The Lancet. 2007;369: 115–122.

52. Elbez-Rubinstein A, Ajzenberg D, Dardé M-L, Cohen R, Dumètre A, Yera H, et al. Congenital toxoplasmosis and reinfection during pregnancy: case report, strain characterization, experimental model of reinfection, and review. J Infect Dis. 2009;199: 280–285. doi:10.1086/595793

53. Dardé ML. Biodiversity in Toxoplasma gondii. Toxoplasma gondii. 1996; 27–41.

54. Galal L, Mercier A, Yera H, de-la-Torre A. Toxoplasmosis in South America: a hybridization-driven disease burden? Trends in Parasitology. 2025;41: 1086–1089. doi:10.1016/j.pt.2025.10.002

55. Haboub M, Mercier A, Durieux M-F, Passebosc-Faure K, Nahri I, Bobelna B, et al. Congenital toxoplasmosis in Martinique: Clinical features and Caribbean genotypes in a comparative study with mainland France. International Journal of Infectious Diseases. 2026. doi:10.1016/j.ijid.2026.108411

56. Leroy J, Houzé S, Dardé M-L, Yéra H, Rossi B, Delhaes L, et al. Severe toxoplasmosis imported from tropical Africa in immunocompetent patients: A case series. Travel Medicine and Infectious Disease. 2020;35: 101509.

57. Fekkar A, Ajzenberg D, Bodaghi B, Touafek F, Le Hoang P, Delmas J, et al. Direct genotyping of Toxoplasma gondii in ocular fluid samples from 20 patients with ocular toxoplasmosis: predominance of type II in France. J Clin Microbiol. 2011;49: 1513–1517. doi:10.1128/JCM.02196-10

58. de-la-Torre A, Sauer A, Pfaff AW, Bourcier T, Brunet J, Speeg-Schatz C, et al. Severe South American ocular toxoplasmosis is associated with decreased Ifn-γ/Il-17a and increased Il-6/Il-13 intraocular levels. PLoS Negl Trop Dis. 2013;7: e2541. doi:10.1371/journal.pntd.0002541

