## Supplementary material for "*Toxoplasma gondii* genotyping in 2,467 human toxoplasmosis cases from France and other geographical origins: genetic diversity and clinical associations": S2-S9 tables

**S2 Table. Comparison of cases with and without a complete 15-marker genotype (n = 2,467)**

|  | **All Cases, n=2,467 (%)** | **With a complete genotype, n=1,269 (%)** | **Complete 15 MS genotyping among all cases of this category, %** |
| --- | --- | --- | --- |
| **CLINICAL CATEGORY** | | | |
| **CT** | 1,377 (55.8) | 955 (75.3) | 69.4 |
| **ATID** | 629 (25.5) | 238 (18.8) | 37.8 |
| **ATIC** | 97 (3.9) | 42 (3.3) | 43.3 |
| **OT** | 325 (13.2) | 23 (1.8) | 7.1 |
| **no CT** | 16 (0.6) | 9 (0.7) | 56.3 |
| **no data** | 23 (0.9) | 2 (0.2) | 8.7 |
| **TYPE OF SAMPLE** | | | |
| **isolated strain** | 1,062 (43.0) | 927 (73.0) | 87.3 |
| **primary clinical sample** | 1,405 (57.0) | 342 (27.0) | 24.3 |
| **PERIOD OF SAMPLING** | | | |
| **Before 2006** | 433 (17.6) | 308 (24.3) | 71.1 |
| **2006–2009** | 491 (19.9) | 236 (18.6) | 48.1 |
| **2010–2015** | 775 (31.4) | 451 (35.5) | 58.2 |
| **2016–2023** | 709 (28.7) | 267 (21.0) | 37.7 |
| **Year not documented** | 59 (2.4) | 7 (0.6) | 11.9 |
| **GEOGRAPHIC ORIGIN (place of acquisition of infection)** | | | |
| **Europe / Central Asia** | 1,739 (70.5) | 1,102 (86.8) | 63.4 |
| **East Asia / Pacific** | 15 (0.6) | 9 (0.7) | 60.0 |
| **LATAM / Caribbean** | 131 (5.3) | 70 (5.5) | 53.4 |
| **MENA** | 5 (0.2) | 1 (0.1) | 0.2 |
| **South Asia** | 1 (0.1) | 0 (0.0) | 0.0 |
| **SSA** | 87 (3.5) | 55 (4.3) | 63.2 |
| **Undefined** | 489 (19.8) | 32 (2.5) | 6.5 |
| **CLINICAL FORM, CONGENITAL TOXOPLASMOSIS** | | | |
| **Infraclinic** | 918 (37.2) | 645 (50.8) | 70.3 |
| **Mild** | 76 (3.1) | 58 (4.6) | 76.3 |
| **Severe** | 195 (7.9) | 151 (11.9) | 77.4 |
| **Severe ocular** | 24 (1.0) | 23 (1.8) | 95.8 |
| **No or insufficient data** | 164 (6.6) | 78 (6.1) | 47.6 |
| **CLINICAL FORM, ACQUIRED TOXOPLASMOSIS IN IMMUNODEFICIENT PATIENTS** | | | |
| **Cerebral** | 284 (11.5) | 99 (7.8) | 34.9 |
| **Disseminated** | 276 (11.2) | 126 (10.0) | 45.7 |
| **Paucisymptomatic or asymptomatic** | 62 (2.5) | 13 (1.0) | 21.0 |
| **No or insufficient data** | 7 (0.3) | 0 (0.0) | 0.0 |
| **CLINICAL FORM, ACQUIRED TOXOPLASMOSIS IN IMMUNOCOMPETENT PATIENTS** | | | |
| **Paucisymptomatic** | 34 (1.4) | 9 (0.7) | 26.5 |
| **Severe** | 59 (2.4) | 33 (2.6) | 55.9 |
| **No or insufficient data** | 4 (0.2) | 0 (0.0) | 0.0 |
| **CLINICAL FORM, ISOLATED OCULAR TOXOPLASMOSIS** | | | |
| **Non-Severe** | 34 (1.4) | 4 (0.3) | 11.8 |
| **Severe** | 148 (6.0) | 15 (1.2) | 10.1 |
| **No or insufficient data** | 143 (5.8) | 4 (0.3) | 2.8 |

*Legend: CT: congenital toxoplasmosis; ATIC: systemic acquired toxoplasmosis in immunocompetent patients; ATID: systemic or localized acquired toxoplasmosis in immunodeficient patients; OT: isolated ocular toxoplasmosis; no CT: congenital toxoplasmosis initially suspected but ultimately excluded after follow-up of the child; LATAM: Latin America; MENA: Middle East and North Africa; SSA: Sub-Saharan Africa. Percentages are column percentages, calculated within each genetic group. Geographic origin refers to the place of acquisition of infection; the country of residence or origin was recorded separately and is not shown here.*

*Among congenital cases with documented clinical data, severe or severe ocular forms accounted for 19.7% (173/877) of successfully genotyped cases versus 13.4% (45/336) of cases without a complete genotype (OR 1.59; 95% CI 1.10–2.32; p = 0.010).*

**S3 Table. Description of *Toxoplasma gondii* genotypes (n = 1,269)**

|  | **Africa 1 N = 34** | **Africa 2 N = 5** | **Africa 4 N = 6** | **Amazonian N = 16** | **Caribbean 1 N = 9** | **Caribbean 2 N = 4** | **Caribbean 3 N = 6** | **Haplogroup 16 N = 10** | **Type I N = 5** | **Type II N = 1,048** | **Type III N = 38** | **Mascarene 1 N = 4** | **Polynesian 1 N = 5** | **Unclassified N = 79** | **Total N = 1,269** |
| --- | --- | --- | --- | --- | --- | --- | --- | --- | --- | --- | --- | --- | --- | --- | --- |
| **MAIN CATEGORIES** | | | | | | | | | | | | | | | |
| **CT** | 6 (17.6) | 0 (0.0) | 1 (16.7) | 2 (12.5) | 2 (22.2) | 4 (100.0) | 3 (50.0) | 7 (70.0) | 2 (40.0) | 869 (82.9) | 21 (55.3) | 2 (50.0) | 5 (100.0) | 31 (39.2) | 955 (75.3) |
| **ATID** | 25 (73.5) | 5 (100.0) | 5 (83.3) | 0 (0.0) | 6 (66.7) | 0 (0.0) | 3 (50.0) | 1 (10.0) | 0 (0.0) | 156 (14.9) | 17 (44.7) | 1 (25.0) | 0 (0.0) | 19 (24.1) | 238 (18.8) |
| **ATIC** | 1 (2.9) | 0 (0.0) | 0 (0.0) | 14 (87.5) | 0 (0.0) | 0 (0.0) | 0 (0.0) | 1 (10.0) | 0 (0.0) | 3 (0.3) | 0 (0.0) | 0 (0.0) | 0 (0.0) | 23 (29.1) | 42 (3.3) |
| **OT** | 2 (5.9) | 0 (0.0) | 0 (0.0) | 0 (0.0) | 1 (11.1) | 0 (0.0) | 0 (0.0) | 1 (10.0) | 0 (0.0) | 14 (1.3) | 0 (0.0) | 1 (25.0) | 0 (0.0) | 4 (5.1) | 23 (1.8) |
| **no CT** | 0 (0.0) | 0 (0.0) | 0 (0.0) | 0 (0.0) | 0 (0.0) | 0 (0.0) | 0 (0.0) | 0 (0.0) | 3 (60.0) | 6 (0.6) | 0 (0.0) | 0 (0.0) | 0 (0.0) | 0 (0.0) | 9 (0.7) |
| **no data** | 0 (0.0) | 0 (0.0) | 0 (0.0) | 0 (0.0) | 0 (0.0) | 0 (0.0) | 0 (0.0) | 0 (0.0) | 0 (0.0) | 0 (0.0) | 0 (0.0) | 0 (0.0) | 0 (0.0) | 2 (2.5) | 2 (0.2) |
| **GEOGRAPHIC ORIGIN (place of acquisition of infection)** | | | | | | | | | | | | | | | |
| **East Asia / Pacific** | 0 (0.0) | 0 (0.0) | 0 (0.0) | 0 (0.0) | 0 (0.0) | 0 (0.0) | 0 (0.0) | 0 (0.0) | 0 (0.0) | 1 (0.1) | 1 (2.6) | 0 (0.0) | 5 (100.0) | 2 (2.5) | 9 (0.7) |
| **Europe / Central Asia** | 2 (5.9) | 0 (0.0) | 2 (33.3) | 0 (0.0) | 3 (33.3) | 0 (0.0) | 0 (0.0) | 10 (100.0) | 5 (100.0) | 1,028 (98.1) | 25 (65.8) | 0 (0.0) | 0 (0.0) | 27 (34.2) | 1,102 (86.8) |
| **LATAM / Caribbean** | 0 (0.0) | 0 (0.0) | 0 (0.0) | 16 (100.0) | 6 (66.7) | 4 (100.0) | 5 (83.3) | 0 (0.0) | 0 (0.0) | 0 (0.0) | 3 (7.9) | 0 (0.0) | 0 (0.0) | 36 (45.6) | 70 (5.5) |
| **MENA** | 0 (0.0) | 0 (0.0) | 1 (16.7) | 0 (0.0) | 0 (0.0) | 0 (0.0) | 0 (0.0) | 0 (0.0) | 0 (0.0) | 0 (0.0) | 0 (0.0) | 0 (0.0) | 0 (0.0) | 0 (0.0) | 1 (0.1) |
| **SSA** | 30 (88.2) | 5 (100.0) | 3 (50.0) | 0 (0.0) | 0 (0.0) | 0 (0.0) | 0 (0.0) | 0 (0.0) | 0 (0.0) | 0 (0.0) | 7 (18.4) | 3 (75.0) | 0 (0.0) | 7 (8.9) | 55 (4.3) |
| **Undefined** | 2 (5.9) | 0 (0.0) | 0 (0.0) | 0 (0.0) | 0 (0.0) | 0 (0.0) | 1 (16.7) | 0 (0.0) | 0 (0.0) | 19 (1.8) | 2 (5.3) | 1 (25.0) | 0 (0.0) | 7 (8.9) | 32 (2.5) |

*Legend: CT: congenital toxoplasmosis; ATIC: systemic acquired toxoplasmosis in immunocompetent patients; ATID: systemic or localized acquired toxoplasmosis in immunodeficient patients; OT: isolated ocular toxoplasmosis; no CT: congenital toxoplasmosis initially suspected but ultimately excluded after follow-up of the child; LATAM: Latin America; MENA: Middle East and North Africa; SSA: Sub-Saharan Africa. Percentages are column percentages, calculated within each genetic group. Geographic origin refers to the place of acquisition of infection; the country of residence or origin was recorded separately and is not shown here.*

**S4 Table. Description of cases in which congenital toxoplasmosis was suspected but ultimately excluded (no CT) (n = 9)**

|  | **Type I N = 3** | **Type II N = 6** | **Total N = 9** |
| --- | --- | --- | --- |
| **PREGNANCY TRIMESTERS** | | | |
| **1** | 1 (33.3) | 1 (16.7) | 2 (22.2) |
| **1–2** | 0 (0.0) | 0 (0.0) | 0 (0.0) |
| **2** | 1 (33.3) | 2 (33.3) | 3 (33.3) |
| **2–3** | 0 (0.0) | 0 (0.0) | 0 (0.0) |
| **3** | 0 (0.0) | 1 (16.7) | 1 (11.1) |
| **Before conception / periconceptional** | 0 (0.0) | 1 (16.7) | 1 (11.1) |
| **Maternal reactivation or reinfection** | 0 (0.0) | 0 (0.0) | 0 (0.0) |
| **Not determined** | 1 (33.3) | 1 (16.7) | 2 (22.2) |
| **GEOGRAPHIC ORIGIN** | | | |
| **East Asia / Pacific** | 0 (0.0) | 0 (0.0) | 0 (0.0) |
| **Europe / Central Asia** | 3 (100.0) | 6 (100.0) | 9 (100.0) |
| **LATAM / Caribbean** | 0 (0.0) | 0 (0.0) | 0 (0.0) |
| **MENA** | 0 (0.0) | 0 (0.0) | 0 (0.0) |
| **SSA** | 0 (0.0) | 0 (0.0) | 0 (0.0) |
| **Undefined** | 0 (0.0) | 0 (0.0) | 0 (0.0) |

*Legend: LATAM: Latin America; SSA: Sub-Saharan Africa.*

**S5 Table. Description of congenital toxoplasmosis (CT) cases (n = 955)**

|  | **Africa 1 N = 6** | **Africa 4 N = 1** | **Amazonian N = 2** | **Caribbean 1 N = 2** | **Caribbean 2 N = 4** | **Caribbean 3 N = 3** | **Haplogroup 16 N = 7** | **Type I N = 2** | **Type II N = 869** | **Type III N = 21** | **Mascarene 1 N = 2** | **Polynesian 1 N = 5** | **Unclassified N = 31** | **Total N = 955** |
| --- | --- | --- | --- | --- | --- | --- | --- | --- | --- | --- | --- | --- | --- | --- |
| **PREGNANCY TRIMESTERS** | | | | | | | | | | | | | | |
| **1** | 1 (16.7) | 0 (0.0) | 1 (50.0) | 0 (0.0) | 0 (0.0) | 1 (33.3) | 1 (14.3) | 2 (100.0) | 107 (12.3) | 1 (4.8) | 0 (0.0) | 2 (40.0) | 7 (22.6) | 123 (12.9) |
| **1–2** | 0 (0.0) | 0 (0.0) | 0 (0.0) | 0 (0.0) | 0 (0.0) | 0 (0.0) | 0 (0.0) | 0 (0.0) | 12 (1.4) | 2 (9.5) | 0 (0.0) | 0 (0.0) | 1 (3.2) | 15 (1.6) |
| **2** | 1 (16.7) | 1 (100.0) | 0 (0.0) | 0 (0.0) | 2 (50.0) | 2 (66.7) | 3 (42.9) | 0 (0.0) | 324 (37.3) | 5 (23.8) | 2 (100.0) | 2 (40.0) | 12 (38.7) | 354 (37.1) |
| **2–3** | 0 (0.0) | 0 (0.0) | 0 (0.0) | 0 (0.0) | 0 (0.0) | 0 (0.0) | 0 (0.0) | 0 (0.0) | 24 (2.8) | 0 (0.0) | 0 (0.0) | 0 (0.0) | 0 (0.0) | 24 (2.5) |
| **3** | 1 (16.7) | 0 (0.0) | 1 (50.0) | 2 (100.0) | 2 (50.0) | 0 (0.0) | 3 (42.9) | 0 (0.0) | 372 (42.8) | 13 (61.9) | 0 (0.0) | 1 (20.0) | 7 (22.6) | 402 (42.1) |
| **Before conception / periconceptional** | 0 (0.0) | 0 (0.0) | 0 (0.0) | 0 (0.0) | 0 (0.0) | 0 (0.0) | 0 (0.0) | 0 (0.0) | 7 (0.8) | 0 (0.0) | 0 (0.0) | 0 (0.0) | 0 (0.0) | 7 (0.7) |
| **Maternal reactivation or reinfection** | 1 (16.7) | 0 (0.0) | 0 (0.0) | 0 (0.0) | 0 (0.0) | 0 (0.0) | 0 (0.0) | 0 (0.0) | 0 (0.0) | 0 (0.0) | 0 (0.0) | 0 (0.0) | 1 (3.2) | 2 (0.2) |
| **Not determined** | 2 (33.3) | 0 (0.0) | 0 (0.0) | 0 (0.0) | 0 (0.0) | 0 (0.0) | 0 (0.0) | 0 (0.0) | 23 (2.6) | 0 (0.0) | 0 (0.0) | 0 (0.0) | 3 (9.7) | 28 (2.9) |
| **CLINICAL FORMS** | | | | | | | | | | | | | | |
| **Infraclinic** | 1 (16.7) | 1 (100.0) | 0 (0.0) | 2 (100.0) | 4 (100.0) | 1 (33.3) | 1 (14.3) | 0 (0.0) | 606 (69.7) | 16 (76.2) | 1 (50.0) | 4 (80.0) | 8 (25.8) | 645 (67.5) |
| **Mild** | 1 (16.7) | 0 (0.0) | 0 (0.0) | 0 (0.0) | 0 (0.0) | 0 (0.0) | 0 (0.0) | 0 (0.0) | 56 (6.4) | 0 (0.0) | 0 (0.0) | 0 (0.0) | 1 (3.2) | 58 (6.1) |
| **Severe** | 4 (66.7) | 0 (0.0) | 2 (100.0) | 0 (0.0) | 0 (0.0) | 2 (66.7) | 4 (57.1) | 2 (100.0) | 115 (13.2) | 2 (9.5) | 0 (0.0) | 1 (20.0) | 19 (61.3) | 151 (15.8) |
| **Severe ocular** | 0 (0.0) | 0 (0.0) | 0 (0.0) | 0 (0.0) | 0 (0.0) | 0 (0.0) | 2 (28.6) | 0 (0.0) | 19 (2.2) | 0 (0.0) | 1 (50.0) | 0 (0.0) | 1 (3.2) | 23 (2.4) |
| **No or insufficient data** | 0 (0.0) | 0 (0.0) | 0 (0.0) | 0 (0.0) | 0 (0.0) | 0 (0.0) | 0 (0.0) | 0 (0.0) | 73 (8.4) | 3 (14.3) | 0 (0.0) | 0 (0.0) | 2 (6.5) | 78 (8.2) |
| **GEOGRAPHIC ORIGIN** | | | | | | | | | | | | | | |
| **East Asia / Pacific** | 0 (0.0) | 0 (0.0) | 0 (0.0) | 0 (0.0) | 0 (0.0) | 0 (0.0) | 0 (0.0) | 0 (0.0) | 1 (0.1) | 1 (4.8) | 0 (0.0) | 5 (100.0) | 2 (6.5) | 9 (0.9) |
| **Europe / Central Asia** | 0 (0.0) | 1 (100.0) | 0 (0.0) | 0 (0.0) | 0 (0.0) | 0 (0.0) | 7 (100.0) | 2 (100.0) | 859 (98.8) | 15 (71.4) | 0 (0.0) | 0 (0.0) | 14 (45.2) | 898 (94.0) |
| **LATAM / Caribbean** | 0 (0.0) | 0 (0.0) | 2 (100.0) | 2 (100.0) | 4 (100.0) | 3 (100.0) | 0 (0.0) | 0 (0.0) | 0 (0.0) | 3 (14.3) | 0 (0.0) | 0 (0.0) | 12 (38.7) | 26 (2.7) |
| **MENA** | 0 (0.0) | 0 (0.0) | 0 (0.0) | 0 (0.0) | 0 (0.0) | 0 (0.0) | 0 (0.0) | 0 (0.0) | 0 (0.0) | 0 (0.0) | 0 (0.0) | 0 (0.0) | 0 (0.0) | 0 (0.0) |
| **SSA** | 6 (100.0) | 0 (0.0) | 0 (0.0) | 0 (0.0) | 0 (0.0) | 0 (0.0) | 0 (0.0) | 0 (0.0) | 0 (0.0) | 1 (4.8) | 1 (50.0) | 0 (0.0) | 3 (9.7) | 11 (1.2) |
| **Undefined** | 0 (0.0) | 0 (0.0) | 0 (0.0) | 0 (0.0) | 0 (0.0) | 0 (0.0) | 0 (0.0) | 0 (0.0) | 9 (1.0) | 1 (4.8) | 1 (50.0) | 0 (0.0) | 0 (0.0) | 11 (1.2) |

*Legend: LATAM: Latin America; SSA: Sub-Saharan Africa. Percentages are column percentages within each genetic group. "Before conception / periconceptional" groups maternal infections acquired before conception or within the periconceptional period; "maternal reactivation or reinfection" refers to congenital infections occurring in women infected before pregnancy.*

**S6 Table. Clinical forms of congenital toxoplasmosis according to the trimester of maternal infection, among type II-associated cases (n = 869)**

| **Clinical form** | **1** | **1–2** | **2** | **2–3** | **3** | **Before conception / periconceptional** | **Maternal reactivation or reinfection** | **Not determined** | **Total** |
| --- | --- | --- | --- | --- | --- | --- | --- | --- | --- |
| **Infraclinic** | 29 | 6 | 222 | 21 | 314 | 2 | 0 | 12 | 606 |
| **Mild** | 6 | 1 | 38 | 0 | 10 | 1 | 0 | 0 | 56 |
| **Severe** | 64 | 5 | 34 | 0 | 4 | 4 | 0 | 4 | 115 |
| **Severe ocular** | 3 | 0 | 7 | 1 | 8 | 0 | 0 | 0 | 19 |
| **No or insufficient data** | 5 | 0 | 23 | 2 | 36 | 0 | 0 | 7 | 73 |
| **Total** | 107 | 12 | 324 | 24 | 372 | 7 | 0 | 23 | 869 |
| **Severe or severe ocular forms, n/documented (%)** | 67/102 (65.7) | 5/12 (41.7) | 41/301 (13.6) | 1/22 (4.5) | 12/336 (3.6) | 4/7 (57.1) | — | 4/16 (25.0) | 134/796 (16.8) |

*Percentages in the last row are calculated among cases with documented clinical data.*

**S7 Table. Description of systemic or localized acquired toxoplasmosis in immunodeficient patients (ATID) cases (n = 238)**

|  | **Africa 1 N = 25** | **Africa 2 N = 5** | **Africa 4 N = 5** | **Caribbean 1 N = 6** | **Caribbean 3 N = 3** | **Haplogroup 16 N = 1** | **Type II N = 156** | **Type III N = 17** | **Mascarene 1 N = 1** | **Unclassified N = 19** | **Total N = 238** |
| --- | --- | --- | --- | --- | --- | --- | --- | --- | --- | --- | --- |
| **CAUSE OF IMMUNODEFICIENCY** | | | | | | | | | | | |
| **HM** | 1 (4.0) | 1 (20.0) | 0 (0.0) | 1 (16.7) | 0 (0.0) | 1 (100.0) | 71 (45.5) | 5 (29.4) | 0 (0.0) | 2 (10.5) | 82 (34.5) |
| **HIV** | 20 (80.0) | 4 (80.0) | 4 (80.0) | 5 (83.3) | 3 (100.0) | 0 (0.0) | 51 (32.7) | 9 (52.9) | 1 (100.0) | 14 (73.7) | 111 (46.6) |
| **SOT** | 3 (12.0) | 0 (0.0) | 1 (20.0) | 0 (0.0) | 0 (0.0) | 0 (0.0) | 20 (12.8) | 3 (17.6) | 0 (0.0) | 1 (5.3) | 28 (11.8) |
| **OI** | 1 (4.0) | 0 (0.0) | 0 (0.0) | 0 (0.0) | 0 (0.0) | 0 (0.0) | 10 (6.4) | 0 (0.0) | 0 (0.0) | 1 (5.3) | 12 (5.0) |
| **No data** | 0 (0.0) | 0 (0.0) | 0 (0.0) | 0 (0.0) | 0 (0.0) | 0 (0.0) | 4 (2.6) | 0 (0.0) | 0 (0.0) | 1 (5.3) | 5 (2.1) |
| **CLINICAL FORMS** | | | | | | | | | | | |
| **Cerebral** | 20 (80.0) | 3 (60.0) | 4 (80.0) | 4 (66.7) | 0 (0.0) | 0 (0.0) | 53 (34.0) | 4 (23.5) | 1 (100.0) | 10 (52.6) | 99 (41.6) |
| **Disseminated** | 5 (20.0) | 2 (40.0) | 1 (20.0) | 2 (33.3) | 3 (100.0) | 1 (100.0) | 90 (57.7) | 13 (76.5) | 0 (0.0) | 9 (47.4) | 126 (52.9) |
| **Paucisymptomatic or asymptomatic** | 0 (0.0) | 0 (0.0) | 0 (0.0) | 0 (0.0) | 0 (0.0) | 0 (0.0) | 13 (8.3) | 0 (0.0) | 0 (0.0) | 0 (0.0) | 13 (5.5) |
| **No or insufficient data** | 0 (0.0) | 0 (0.0) | 0 (0.0) | 0 (0.0) | 0 (0.0) | 0 (0.0) | 0 (0.0) | 0 (0.0) | 0 (0.0) | 0 (0.0) | 0 (0.0) |
| **GEOGRAPHIC ORIGIN** | | | | | | | | | | | |
| **East Asia / Pacific** | 0 (0.0) | 0 (0.0) | 0 (0.0) | 0 (0.0) | 0 (0.0) | 0 (0.0) | 0 (0.0) | 0 (0.0) | 0 (0.0) | 0 (0.0) | 0 (0.0) |
| **Europe / Central Asia** | 1 (4.0) | 0 (0.0) | 1 (20.0) | 3 (50.0) | 0 (0.0) | 1 (100.0) | 146 (93.6) | 10 (58.8) | 0 (0.0) | 6 (31.6) | 168 (70.6) |
| **LATAM / Caribbean** | 0 (0.0) | 0 (0.0) | 0 (0.0) | 3 (50.0) | 2 (66.7) | 0 (0.0) | 0 (0.0) | 0 (0.0) | 0 (0.0) | 5 (26.3) | 10 (4.2) |
| **MENA** | 0 (0.0) | 0 (0.0) | 1 (20.0) | 0 (0.0) | 0 (0.0) | 0 (0.0) | 0 (0.0) | 0 (0.0) | 0 (0.0) | 0 (0.0) | 1 (0.4) |
| **SSA** | 22 (88.0) | 5 (100.0) | 3 (60.0) | 0 (0.0) | 0 (0.0) | 0 (0.0) | 0 (0.0) | 6 (35.3) | 1 (100.0) | 4 (21.1) | 41 (17.2) |
| **Undefined** | 2 (8.0) | 0 (0.0) | 0 (0.0) | 0 (0.0) | 1 (33.3) | 0 (0.0) | 10 (6.4) | 1 (5.9) | 0 (0.0) | 4 (21.1) | 18 (7.6) |

*Legend: HIV: human immunodeficiency virus; HM: haematological malignancies or haematopoietic stem cell transplantation; OI: other causes of immunosuppression (solid cancer, primary or unexplained immunodeficiency, immunosuppressive or cytotoxic treatment, chronic kidney or liver disease); SOT: solid organ transplantation; LATAM: Latin America; MENA: Middle East and North Africa; SSA: Sub-Saharan Africa.*

**S8 Table. Description of systemic acquired toxoplasmosis in immunocompetent patients (ATIC) cases (n = 42)**

|  | **Africa 1 N = 1** | **Amazonian N = 14** | **Haplogroup 16 N = 1** | **Type II N = 3** | **Unclassified N = 23** | **Total N = 42** |
| --- | --- | --- | --- | --- | --- | --- |
| **CLINICAL FORMS** | | | | | | |
| **Paucisymptomatic** | 1 (100.0) | 0 (0.0) | 0 (0.0) | 3 (100.0) | 5 (21.7) | 9 (21.4) |
| **Severe** | 0 (0.0) | 14 (100.0) | 1 (100.0) | 0 (0.0) | 18 (78.3) | 33 (78.6) |
| **No or insufficient data** | 0 (0.0) | 0 (0.0) | 0 (0.0) | 0 (0.0) | 0 (0.0) | 0 (0.0) |
| **GEOGRAPHIC ORIGIN** | | | | | | |
| **East Asia / Pacific** | 0 (0.0) | 0 (0.0) | 0 (0.0) | 0 (0.0) | 0 (0.0) | 0 (0.0) |
| **Europe / Central Asia** | 0 (0.0) | 0 (0.0) | 1 (100.0) | 3 (100.0) | 3 (13.0) | 7 (16.7) |
| **LATAM / Caribbean** | 0 (0.0) | 14 (100.0) | 0 (0.0) | 0 (0.0) | 19 (82.6) | 33 (78.6) |
| **MENA** | 0 (0.0) | 0 (0.0) | 0 (0.0) | 0 (0.0) | 0 (0.0) | 0 (0.0) |
| **SSA** | 1 (100.0) | 0 (0.0) | 0 (0.0) | 0 (0.0) | 0 (0.0) | 1 (2.4) |
| **Undefined** | 0 (0.0) | 0 (0.0) | 0 (0.0) | 0 (0.0) | 1 (4.3) | 1 (2.4) |

*Legend: LATAM: Latin America; SSA: Sub-Saharan Africa. One of these cases is the mother of a congenital case reported in S2 Table (same strain, French Guiana, 2014). Several ATIC strains from French Guiana share identical 15-marker profiles within the same year and correspond to documented common-source outbreaks; these observations are therefore not independent.*

**S9 Table. Description of isolated ocular toxoplasmosis (OT) cases (n = 23)**

|  | **Africa 1 N = 2** | **Caribbean 1 N = 1** | **Haplogroup 16 N = 1** | **Type II N = 14** | **Mascarene 1 N = 1** | **Unclassified N = 4** | **Total N = 23** |
| --- | --- | --- | --- | --- | --- | --- | --- |
| **CLINICAL FORMS** | | | | | | | |
| **Non-severe** | 0 (0.0) | 0 (0.0) | 0 (0.0) | 3 (21.4) | 0 (0.0) | 1 (25.0) | 4 (17.4) |
| **Severe** | 2 (100.0) | 1 (100.0) | 1 (100.0) | 8 (57.1) | 1 (100.0) | 2 (50.0) | 15 (65.2) |
| **No or insufficient data** | 0 (0.0) | 0 (0.0) | 0 (0.0) | 3 (21.4) | 0 (0.0) | 1 (25.0) | 4 (17.4) |
| **IMMUNE STATUS** | | | | | | | |
| **Immunocompetent** | 1 (50.0) | 1 (100.0) | 0 (0.0) | 11 (78.6) | 1 (100.0) | 2 (50.0) | 16 (69.6) |
| **Immunodeficient** | 1 (50.0) | 0 (0.0) | 1 (100.0) | 2 (14.3) | 0 (0.0) | 1 (25.0) | 5 (21.7) |
| **Not documented** | 0 (0.0) | 0 (0.0) | 0 (0.0) | 1 (7.1) | 0 (0.0) | 1 (25.0) | 2 (8.7) |
| **GEOGRAPHIC ORIGIN** | | | | | | | |
| **East Asia / Pacific** | 0 (0.0) | 0 (0.0) | 0 (0.0) | 0 (0.0) | 0 (0.0) | 0 (0.0) | 0 (0.0) |
| **Europe / Central Asia** | 1 (50.0) | 0 (0.0) | 1 (100.0) | 14 (100.0) | 0 (0.0) | 3 (75.0) | 19 (82.6) |
| **LATAM / Caribbean** | 0 (0.0) | 1 (100.0) | 0 (0.0) | 0 (0.0) | 0 (0.0) | 0 (0.0) | 1 (4.3) |
| **MENA** | 0 (0.0) | 0 (0.0) | 0 (0.0) | 0 (0.0) | 0 (0.0) | 0 (0.0) | 0 (0.0) |
| **SSA** | 1 (50.0) | 0 (0.0) | 0 (0.0) | 0 (0.0) | 1 (100.0) | 0 (0.0) | 2 (8.7) |
| **Undefined** | 0 (0.0) | 0 (0.0) | 0 (0.0) | 0 (0.0) | 0 (0.0) | 1 (25.0) | 1 (4.3) |

*Legend: LATAM: Latin America; SSA: Sub-Saharan Africa. Severity was defined by the presence of at least one of the following: extensive inflammation (e.g. panuveitis), bilateral involvement, recurrent or reactivated disease, or incomplete clinical improvement. In practice, most severe cases were classified as such on the basis of recurrent or reactivated disease; duration of follow-up was not documented.*
